# Transcriptome-Wide Root Causal Inference

**DOI:** 10.1101/2024.07.22.24310837

**Authors:** Eric V. Strobl, Eric R. Gamazon

**Affiliations:** University of Pittsburgh; Vanderbilt University Medical Center

## Abstract

Root causal genes correspond to the first gene expression levels perturbed during pathogenesis by genetic or non-genetic factors. Targeting root causal genes has the potential to alleviate disease entirely by eliminating pathology near its onset. No existing algorithm discovers root causal genes from observational data alone. We therefore propose the Transcriptome-Wide Root Causal Inference (TWRCI) algorithm that identifies root causal genes and their causal graph using a combination of genetic variant and unperturbed bulk RNA sequencing data. TWRCI uses a novel competitive regression procedure to annotate cis and trans-genetic variants to the gene expression levels they directly cause. The algorithm simultaneously recovers a causal ordering of the expression levels to pinpoint the underlying causal graph and estimate root causal effects. TWRCI outperforms alternative approaches across a diverse group of metrics by directly targeting root causal genes while accounting for distal relations, linkage disequilibrium, patient heterogeneity and widespread pleiotropy. We demonstrate the algorithm by uncovering the root causal mechanisms of two complex diseases, which we confirm by replication using independent genome-wide summary statistics.

## 1 Introduction

Genetic and non-genetic factors can influence gene expression levels to ultimately cause disease. Root causal gene expression levels – or *root causal genes* for short – correspond to the *initial* changes to *gene expression* that ultimately generate disease as a downstream effect^1^. Root causal genes differ from core genes that directly cause the phenotype and thus lie at the end, rather than at the beginning, of pathogenesis^2^. Root causal genes also generalize driver genes that only account for the effects of somatic mutations primarily in protein coding sequences in cancer^3^.

Discovering root causal genes is critical towards identifying drug targets that modify disease near its pathogenic onset and thus mitigate downstream pathogenesis in its entirety^4^. The problem is complicated by the existence of complex disease, where the causal effects of the root causal genes may differ between patients even within the same diagnostic category. However, the recently defined *omnigenic root causal model* posits that only a few root causal genes affect nearly all downstream genes to initiate the vast majority of pathology in each patient^1^. We thus more specifically seek to identify *personalized* root causal genes specific to any given individual.

Only one existing algorithm accurately identifies personalized root causal genes^1^, but the algorithm requires access to genome-wide Perturb-seq data, or high throughput perturbations with single cell RNA sequencing readout^5, 6^. Perturb-seq is currently expensive and difficult to obtain in many cell types. We instead seek a method that can uncover personalized root causal genes directly from widespread observational (or non-experimental) datasets.

We make the following contributions in this paper:

1. We introduce the conditional root causal effect (CRCE) that measures the causal effect of the genetic and non-genetic factors, which directly affect a gene expression level, on the phenotype.
2. We propose a novel strategy called Competitive Regression that provably annotates both cis and trans-genetic variants to the gene expression level or phenotype they directly cause without conservative significance testing.
3. We create an algorithm called Trascriptome-Wide Root Causal Inference (TWRCI) that uses the annotations to reconstruct a personalized causal graph summarizing the CRCEs of gene expression levels from a combination of genetic variant and bulk RNA sequencing observational data.
4. We show with confirmatory replication that TWRCI identifies only a few root causal genes that accurately distinguish subgroups of patients even in complex diseases – consistent with the omnigenic root causal model.

We provide an example of the output of TWRCI in Figure 1. TWRCI annotates both cis and trans genetic variants to the expression level or phenotype they *directly* cause. We prove that the direct causal annotations allow the algorithm to uniquely reconstruct the causal graph between the gene expression levels that cause the phenotype as well as estimate their CRCEs. The algorithm summarizes the CRCEs in the graph by weighing and color-coding each vertex, where vertex size correlates with magnitude, green induces disease and red prevents disease. TWRCI thus provides a succinct summary of root causal genes and their root causal effect sizes specific to a given patient using observational data alone. TWRCI outperforms combinations of existing algorithms across all subtasks: annotation, graph reconstruction and CRCE estimation. No existing algorithm performs all subtasks simultaneously.

**Figure 1.**
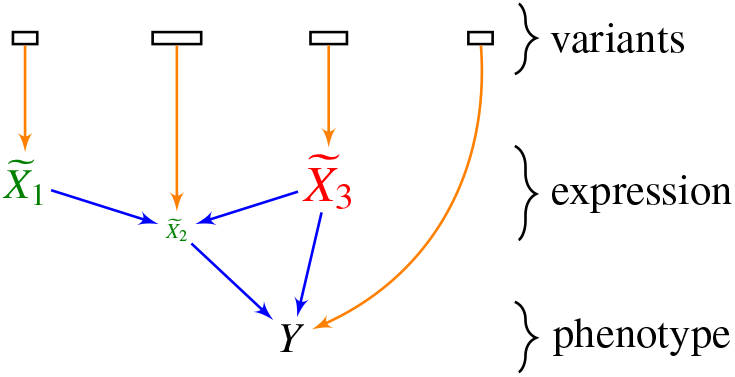
Toy example of a root causal mechanism inferred by TWRCI for a specific patient. Rectangles denote sets of genetic variants, potentially in linkage disequilibrium even between sets. Each set of variants directly causes a gene expression level in 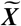 or the phenotype *Y*. Larger lettered vertices denote larger CRCE magnitudes and colors refer to their direction – green is a positive CRCE and red is negative. TWRCI simultaneously annotates, reconstructs and estimates the CRCEs.

## 2 Results

### 2.1 Overview of TWRCI

#### 2.1.1 Setup

We seek to identify not just causal but *root causal* genes. We must therefore carefully define the generative process. We consider a set of variants ***S***, the transcriptome 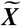 and the phenotype *Y*. We represent the generative causal process using a directed graph like in Figure 2 (a), where the variants cause the transcriptome, and the transcriptome causes the phenotype. Directed edges denote direct causal relations between variables. In practice, the sets ***S*** and 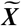 contain millions and thousands of variables, respectively. As described in Methods 4.2, we cannot measure the values of 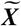 exactly using RNA sequencing but instead measure values ***X*** corrupted by Poisson measurement error and batch effects.

**Figure 2.**
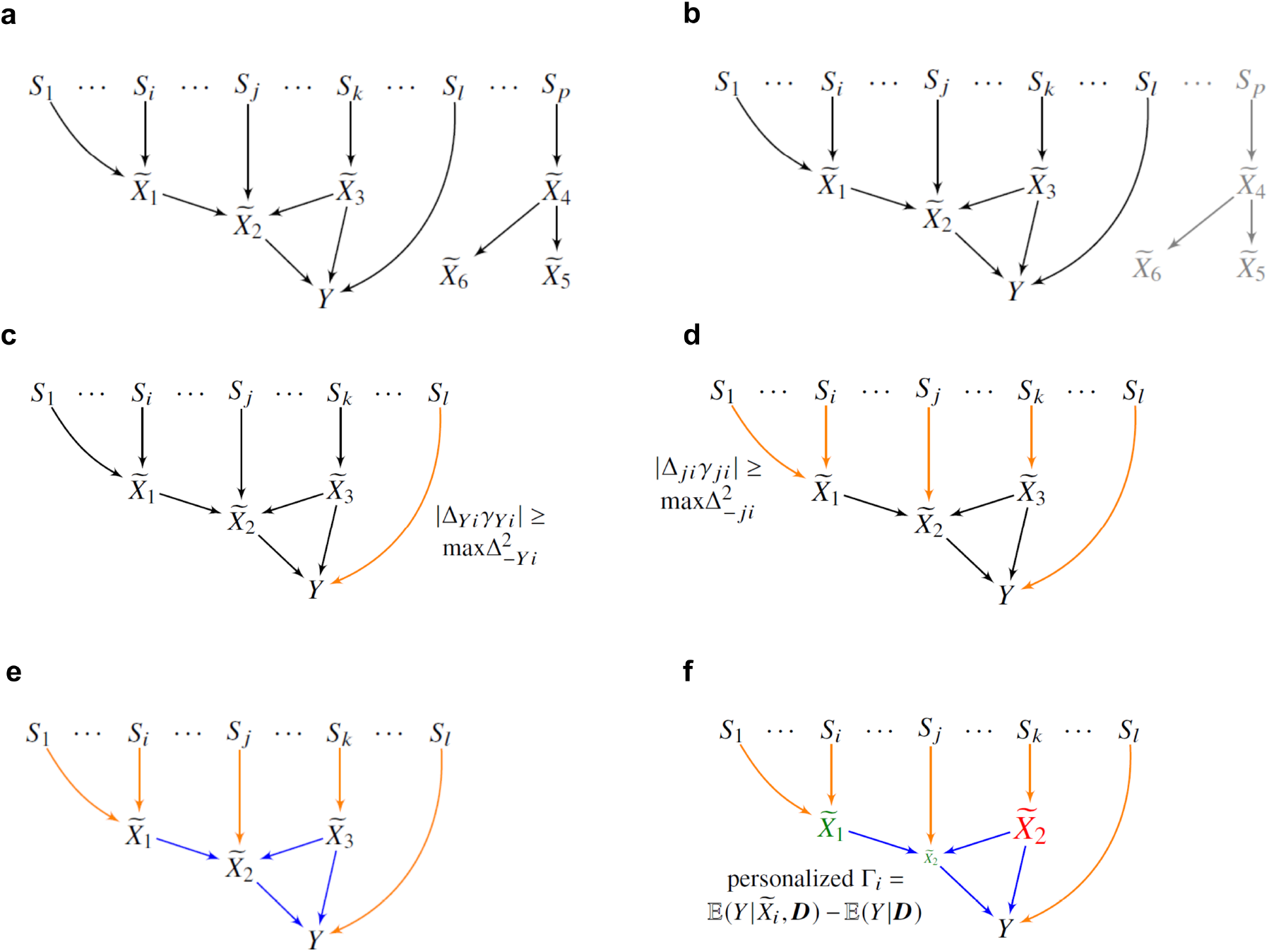
Overview of the TWRCI algorithm. (a) We redraw Figure 1 in more detail. We do not have access to the underlying causal graph in practice. (b) TWRCI first performs variable selection by only keeping variants and gene expression levels correlated with *Y* (and their common cause confounders) as shown in black. (c) The algorithm then uses Competitive Regression to find the variants that directly cause *Y* in orange. (d) TWRCI iteratively repeats Competitive Regression for each gene expression level as well – again shown in orange. (e) The algorithm next performs causal discovery to identify the causal relations between the gene expression levels and the phenotype in blue. (f) Finally, TWRCI weighs each vertex 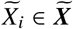 by the magnitude of Γ_*i*_ and color codes the vertex by its direction (green is positive, red is negative). TWRCI thus ultimately recovers a causal graph like the one shown in Figure 1.

#### 2.1.2 Variable Selection

Simultaneously handling millions of variants and thousands of gene expression levels currently requires expensive computational resources. Moreover, most variants and gene expression levels do not inform the discovery of root causal genes for a particular phenotype *Y*. The Transcriptome-Wide Root Causal Inference (TWRCI) thus first performs variable selection by eliminating variants and gene expression levels unnecessary for root causal inference.

TWRCI first identifies variants ***T*** ⊆ ***S*** associated with *Y* using widely available summary statistics at a liberal *α* threshold, such as 5e-5, in order to capture many causal variants. The algorithm then uses individual-level data – where each individual has variant data, bulk gene expression data from the relevant tissue and phenotype data (variant-expression-phenotype) – to learn a regression model predicting ***X*** from ***T***. TWRCI identifies the subset of expression levels 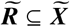 that it can predict better than chance. We refer the reader to Methods 4.4.2 for details on the discovery of additional nuisance variables required to mitigate confounding. We prove that 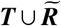 retains all of the causes of *Y* in 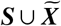.

#### 2.1.3 Annotation by Competitive Regression

We want to annotate both cis and trans-variants to the gene expression level that they directly cause in 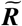. We also want to annotate variants to the phenotype *Y* in order to account for horizontal pleiotropy, where variants bypass 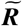 and directly cause *Y*. TWRCI achieves both of these feats through a novel process called *Competitive Regression*.

TWRCI accounts for horizontal pleiotropy by applying Competitive Regression to the phenotype. We do not restrict the theoretical results detailed in Methods to linear models, but linear models trained on genotype data currently exhibit competitive performance^7, 8^. TWRCI therefore trains debiased linear ridge regression models^9^ predicting *Y* from 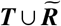 without requiring Gaussian distributions; let *γ*_*Yi*_ refer to the coefficient for *T*_*i*_ in the regression model. Similarly, let Δ_−*Yi*_ correspond to the matrix of coefficients for *T*_*i*_ in the regression models predicting 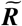 (but not *Y*) from ***T***; notice that we have not conditioned on the gene expression levels in this case. If *T*_*i*_ *directly* causes *Y*, then it will predict *Y* given ***T*** \ *T*_*i*_ and given 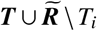 (i.e., Δ_*Yi*_ and *γ*_*Yi*_ will both be non-zero), but *T*_*i*_ will not predict any gene expression level given ***T*** \ *T*_*i*_ (i.e., 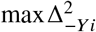 will be zero). We also prove the converse direction in Methods 4.4.4. TWRCI therefore annotates *T*_*i*_ to *Y*, if |Δ_*Yi*_*γ*_*Yi*_ | deviates away from zero even after conditioning on gene expression so that 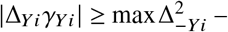 i.e., Δ_*Yi*_*γ*_*Yi*_ “beats” 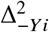 in a competitive process (Figure 2 (c)).

We prove in Methods 4.4.3 that Competitive Regression successfully recovers the direct causes of *Y*, denoted by ***S***_*Y*_ ⊆ ***T***, so long as *Y* is a sink vertex that does not cause any other variable. We also require analogues of two standard assumptions used in instrumental variable analysis: relevance and exchangeability^10^. In this paper, *relevance* means that at least one variant in ***T*** directly causes each gene expression level in 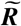; the assumption usually holds because ***T*** contains orders of magnitude more variants than entries in 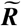. On the other hand, *exchangeability* assumes that ***T*** and other sets of direct causal variants not in ***T*** share no latent confounders (details in Methods 4.4.2); this assumption holds approximately due to the weak causal relations emanating from variants to gene expression and the phenotype. Exchangeability also weakens as ***T*** grows larger.

We further show that Competitive Regression can recover ***S***_*i*_ ⊆ ***T***, or the direct causes of 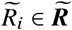 in ***T***, when 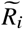 causes *Y* and turns into a sink vertex after removing *Y* from consideration (Methods 4.4.4). As a result, TWRCI removes *Y* and appends it to the empty ordered set ***K*** to ensure that some 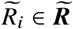 is now a sink vertex. We introduce a statistical criterion in Methods 4.4.4 that allows TWRCI to find the sink vertex 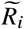 after removing *Y*. TWRCI then annotates ***S***_*i*_ to 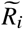 again using Competitive Regression (Figure 2 (d)), removes *R*_*i*_ from ***R*** and appends *R*_*i*_ to the front of ***K***. The algorithm iterates until it has removed all variables from ***R*** ⋃ *Y* and placed them into the causal order ***K***.

#### 2.1.4 Causal Discovery and CRCE Estimation

Annotation only elucidates the direct causal relations from variants to gene expression, but it does not recover the causal relations between gene expression or the causal relations from gene expression to the phenotype. We want TWRCI to recover the *entire* biological mechanism from variants all the way to the phenotype.

TWRCI thus subsequently runs a causal discovery algorithm with the causal order ***K*** to uniquely identify the causal graph over 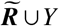 (Figure 2 (e)). The algorithm also estimates the personalized or *conditional root causal effect* (CRCE) of gene expression levels that cause *Y* :

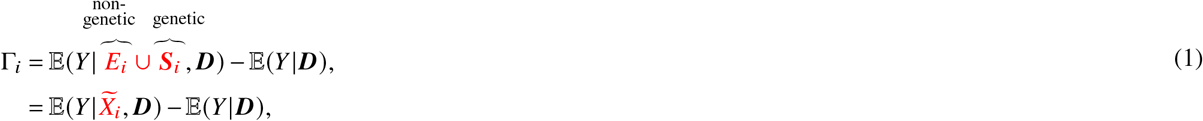

where we choose 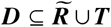 carefully to ensure that the second equality holds (Methods 4.3). The CRCE Γ_*i*_ of 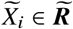 thus measures the causal effect of the genetic factors ***S***_*i*_ and the non-genetic factors *E*_*i*_ on *Y* that perturb 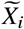 first. The CRCE values differ between patients, so TWRCI can recover different causal graphs by weighing each vertex according to the patient-specific CRCE values Γ = *γ* (Figure 2 (f)). The gene 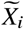 is a *personalized root causal gene* if |*γ*_*i*_| > 0. The *omnigenic root causal model* posits that |*γ*| ≫ 0 for only a small subset of genes in each patient even in complex disease.

### 2.2 TWRCI accurately annotates, reconstructs and estimates in silico

No existing algorithm recovers personalized root causal genes from observational data alone. However, existing algorithms can annotate variants using different criteria and reconstruct causal graphs from observational data. We therefore compared TWRCI against state of the art algorithms in annotation and causal graph reconstruction using 100 semi-synthetic datasets with real variant data but simulated gene expression and phenotype data (Methods 4.6).

Many different annotation methods exist with different objectives. Most methods nevertheless annotate variants by at least considering proximity to the transcription start site (TSS), with the hope that variants near the TSS of a gene will *directly* affect that gene’s expression level; for example, a variant in the exonic region of a gene may compromise its mRNA stability, while a variant in the promoter region may affect its transcription rate. We thus compare a diverse range of methods in *direct causal* annotation, or assigning variants to the gene expression levels they directly cause. This criterion accommodates other annotation objectives from a mathematical perspective as well – solving direct causation automatically solves causation, colocalization and correlation as progressively more relaxed cases. We in particular compare nearest TSS, a one mega-base cis-window^1^, the causal transcriptome-wide association study (cTWAS)^11^, the maximally correlated gene within the cis-window (cis-eQTL)^12^, colocalization with approximate Bayes factors (ABF)^13^, and colocalization with Sum of SIngle Effects model (SuSIE)^14^. We then performed causal graph reconstruction using SIGNET^15, 16^, RCI^17^, GRCI^18^ and the PC algorithm^19, 20^. We evaluated TWRCI against all combinations of annotation and graph reconstruction methods. See Methods 4.5 and 4.8 for a detailed description of comparator algorithms and evaluation metrics, respectively. All statements about empirical results mentioned below hold at a Bonferroni corrected threshold of 0.05 divided by the number of comparator algorithms according to two-sided paired t-tests.

We first summarize the accuracy results for annotation of direct causes only. All existing annotation algorithms utilize heuristics such as location, correlation or colocalization to infer causality. Only TWRCI provably identifies the direct causes of each gene expression level (Theorem 1 in Methods 4.4.6). Empirical results corroborate this theoretical conclusion. TWRCI achieved the highest accuracy as assessed by Matthew’s correlation coefficient (MCC) to the true direct causal variants of each gene expression level and phenotype (Figure 3 (a) left). The algorithm also ranked the ground truth direct causal variants the highest by assigning the ground truth causal variants larger regression coefficient magnitudes than non-causal variants (Figure 3 (a) right). Both TWRCI and cTWAS account for horizontal pleiotropy, but TWRCI again outperformed cTWAS even when we only compared the true and inferred variants that directly cause the phenotype using MCC and the normalized rank (Figure 3 (b)). We conclude that TWRCI annotated the genetic variants to their direct effects most accurately.

**Figure 3.**
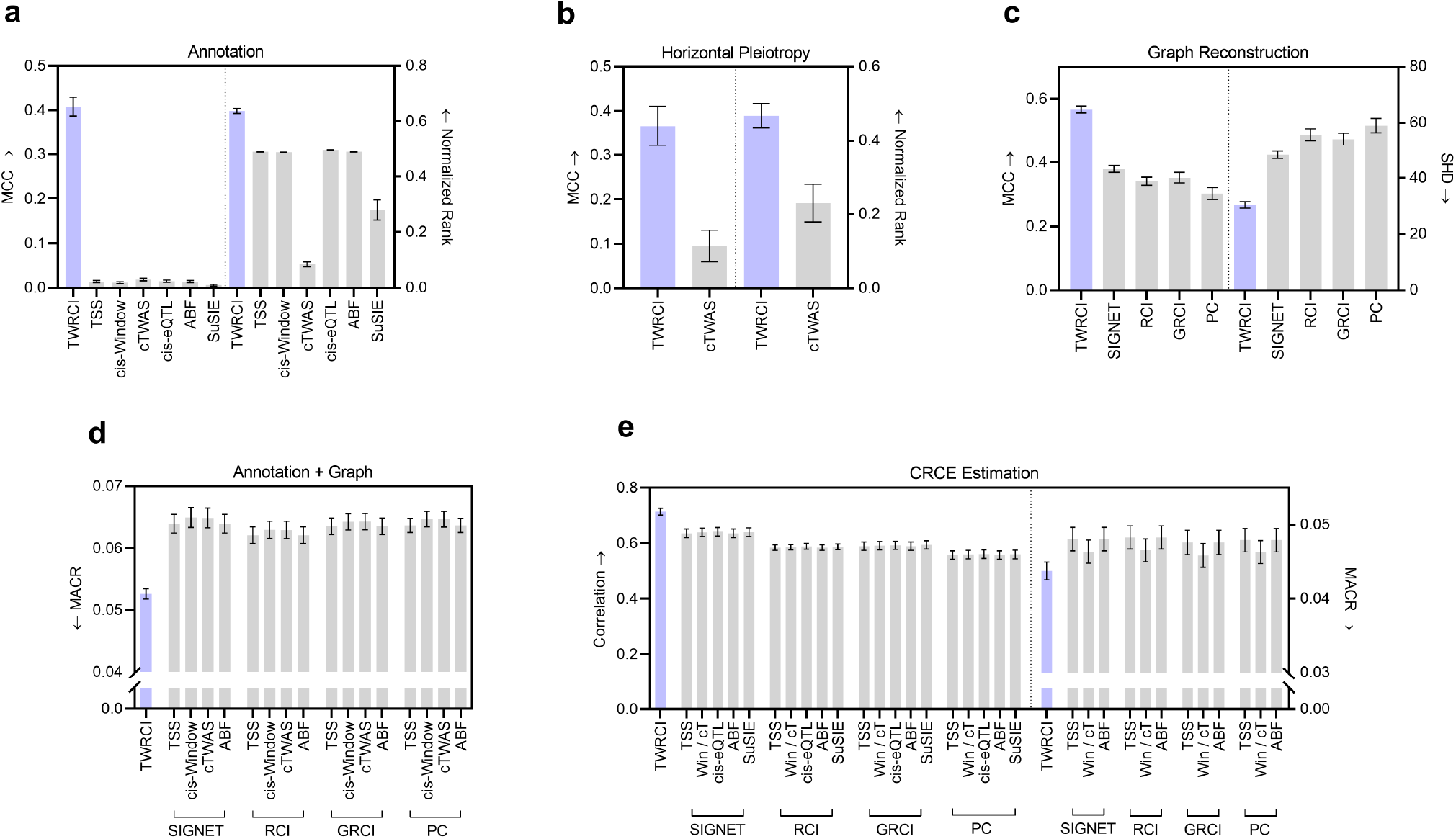
Semi-synthetic data results in terms of (a) direct causal annotation, (b) annotation focused on horizontal pleiotropy only, (c) graph reconstruction, (d) combined annotation and graph reconstruction, and (e) CRCE estimation accuracy. Four of the graphs summarize two evaluation metrics. Arrows near the y-axis denote whether a higher (upward arrow) or a lower (downward arrow) score is better. We do not plot the results of cis-eQTL and SuSIE in (d) and (e) when they exhibit much worse performance. The cis-window and cTWAS algorithms have the exact same CRCE estimates in (e) because accounting for horizontal pleiotropy in cTWAS does not change the conditioning set ***D*** in Equation (1); we thus denote cis-Window and cTWAS as Win/cT for short. TWRCI in purple outperformed all algorithms across all nine evaluation metrics. Error bars correspond to 95% confidence intervals.

We obtained similar results with causal graph reconstruction. TWRCI obtained the best performance according to the highest MCC and the lowest structural hamming distance (SHD) to the ground truth causal graphs (Figure 3 (c)). We then assessed the performance of combined annotation and graph reconstruction using the mean absolute correlation of the residuals (MACR), or the mean absolute correlation between the *indirect* causes of a gene expression level and the residual gene expression level obtained after partialing out the inferred *direct* causes; if an algorithm annotates and reconstructs accurately, then each gene expression level should not correlate with its indirect causes after partialing out its direct causes, so the MACR should attain a small value. TWRCI accordingly achieved the lowest MACR as compared to all possible combinations of existing algorithms (Figure 3 (d)). The cis-eQTL and SuSIE algorithms obtained MACR values greater than 0.3 because many cis-variants did not correlate or colocalize with the expression level of the gene with the nearest TSS; we thus do not plot the results of these algorithms. We conclude that TWRCI used annotations to reconstruct the causal graph most accurately by provably accounting for both cis and trans-variants.

We finally analyzed CRCE estimation accuracy. Computing the CRCE requires access to the inferred annotations and causal graph. We therefore again evaluated TWRCI against all possible combinations of existing algorithms. The CRCE estimates of TWRCI attained the largest correlation to the ground truth CRCE values (Figure 3 (e) left). Further, if an algorithm accurately estimates the components 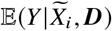 and 𝔼(*Y*|***D***) of the CRCE in Equation (1), then the residual 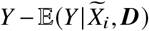 should not correlate with ***S***_*i*_ ⋂ ***T***. TWRCI accordingly obtained the lowest mean absolute correlation of these residuals (MACR) against all combinations of algorithms (Figure 3 (e) right). The cis-eQTL and SuSIE algorithms again attained much worse MACR values above 0.4 because they failed to annotate many causal variants to their gene expression levels. We conclude that TWRCI outperformed existing methods in CRCE estimation. TWRCI therefore annotated, reconstructed and estimated the most accurately according to all nine evaluation criteria. The algorithm also completed within about 3 minutes for each dataset (Supplementary Figure 1).

### 2.3 Chronic and exaggerated immunity in COPD

We next ran the algorithms using summary statistics of a large GWAS of COPD^21^ consisting of 13,530 cases and 454,945 controls of European ancestry. We downloaded individual variant-expression-phenotype data of lung tissue from GTEx^22^ with 96 cases and 415 controls. We also replicated results using an independent GWAS consisting of 4,017 cases and 162,653 controls of East Asian ancestry^21^. COPD is a chronic inflammatory condition of the airways or the alveoli that leads to persistent airflow obstruction^23^. Exposure to respiratory infections or environmental pollutants can also trigger acute on chronic inflammation called COPD exacerbations that worsen the obstruction.

#### 2.3.1 Accuracy

We first compared the accuracy of the algorithms in variant annotation, graph reconstruction and CRCE estimation. We can compute the MACR metrics – representing two of the nine evaluation criteria used in the previous section – with real data. We summarize the MACR for simultaneous variant annotation and graph reconstruction averaged over ten nested cross-validation folds in Figure 4 (a). TWRCI achieved the lowest MACR out of all combinations of algorithms within about 3 minutes (Supplementary Figure 2 (b) and (c)). Performance differed primarily by the annotation method rather than the causal discovery algorithm. Conservative annotation algorithms, such as colocalization by SuSIE, again failed to achieve a low MACR because they frequently failed to annotate at least one variant to every gene expression level. MACR values for CRCE estimation followed a similar pattern (Figure 4 (b)) because accurate annotation and reconstruction enabled accurate downstream CRCE estimation.

**Figure 4.**
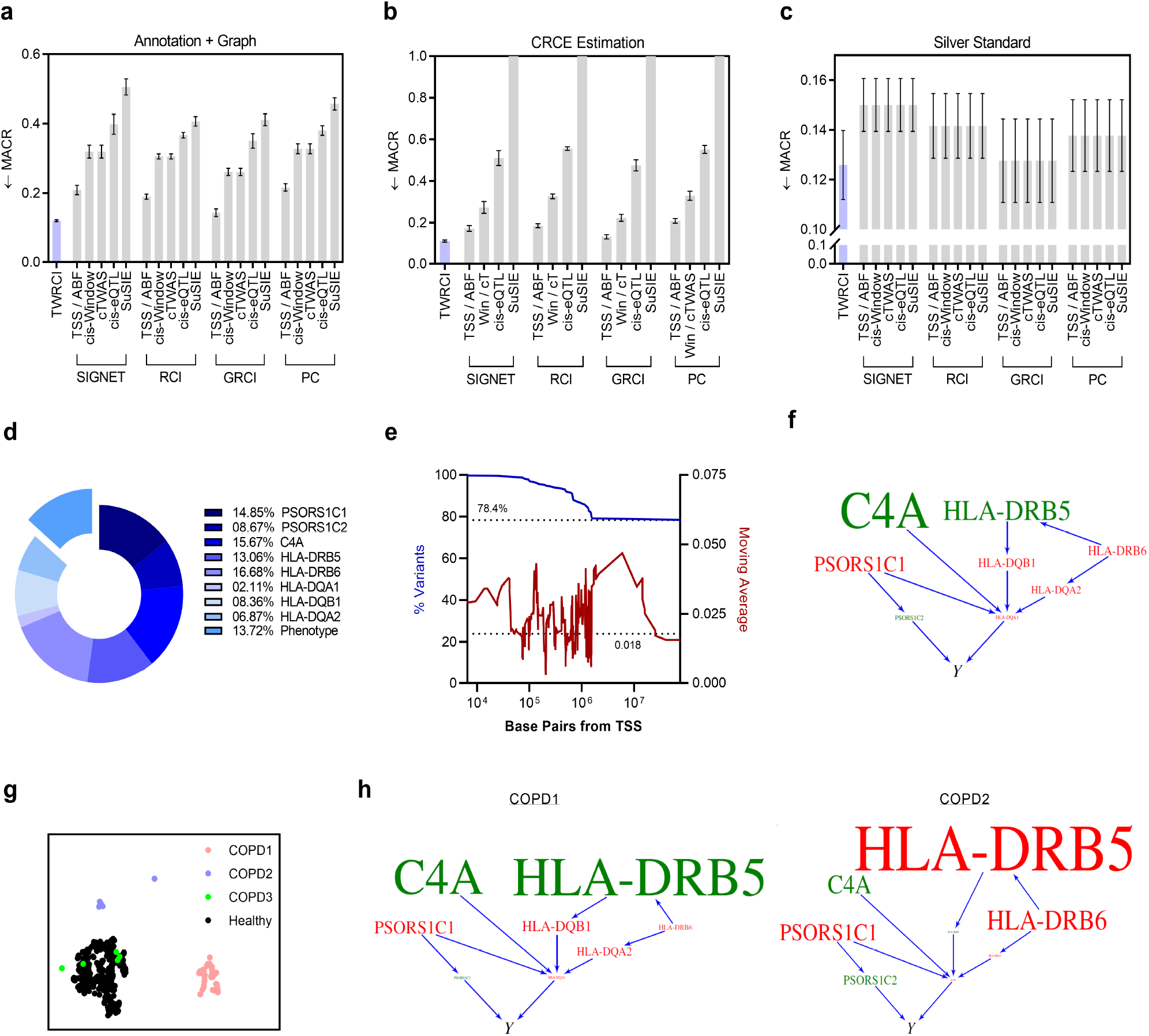
Results for COPD. (a) TWRCI outperformed all other combinations of algorithms in direct causal annotation and graph reconstruction by achieving the lowest MACR; error bars correspond to one standard error of the mean in accordance with the one standard error rule of cross-validation^27^. (b) TWRCI similarly achieved the lowest MACR for CRCE estimation. (c) Silver standard genes exhibited the smallest correlation with the phenotype after partialing out the root causal genes inferred by TWRCI. (d) More than 13% the causal variants exhibited horizontal pleiotropy. TWRCI annotated the remaining causal variants to eight gene expression levels. (e) TWRCI assigned approximately 78% of the causal variants to genes located on different chromosomes. Most causal variants annotated to a gene on the same chromosome fell within a one megabase distance from the TSS (blue, left). The average magnitude of the regression coefficients remained approximately constant with increasing distance from the TSS (red, right); the dotted line again corresponds to variants on different chromosomes. (f) The COPD-wide causal graph revealed multiple MHC class II genes as root causal. (g) UMAP dimensionality reduction revealed two clusters of COPD patients well-separated from the healthy controls. (h) The directed graphs highlighted different root causal genes within each of the two clusters.

We next followed^11, 24^ and downloaded a set of silver standard genes enriched in genes that cause COPD. The KEGG database does not contain a pathway for COPD, so we downloaded the gene set from the DisGeNet database instead (UMLS C0024117, curated)^25, 26^. Many silver standard genes are causal but not *root* causal for COPD. If an algorithm truly identifies root causal genes, then partialing out the root causal genes from all of the downstream non-root causal genes and the phenotype should explain away the vast majority of the causal effect between the non-root causal genes and the phenotype according to the omnigenic root causal model. We therefore computed another MACR metric, the mean absolute correlation between the residuals of the silver standard genes and the residuals of the phenotype after partialing out the inferred root causal genes. TWRCI again obtained the lowest MACR value (Figure 4 (c)). We conclude that TWRCI identified the root causal genes most accurately according to known causal genes in COPD.

#### 2.3.2 Horizontal pleiotropy and trans-variants

We studied the output of TWRCI in detail to gain insight into important issues in computational genomics. Previous studies have implicated the existence of widespread horizontal pleiotropy in many diseases^28^. TWRCI can annotate variants directly to the phenotype, so we can use TWRCI to assess the existence of widespread pleiotropy. The variable selection step of TWRCI identified fourteen gene expression levels surviving false discovery rate (FDR) correction at a liberal 10% threshold; eight of these levels ultimately caused the phenotype, including two psoriasis susceptibility genes, a complement protein and five MHC class II genes. TWRCI annotated 13.7% of the variants that cause COPD directly to the phenotype, despite competition for variants between the phenotype and the eight gene expression levels (Figure 4 (d)). Many variants thus directly cause COPD by bypassing expression. We conclude that TWRCI successfully identified widespread horizontal pleiotropy in COPD. In contrast, cTWAS failed to identify any variants that bypass gene expression because all variants had very small effects on the phenotype, especially after accounting for gene expression; as a result, no variants ultimately had a posterior inclusion probability greater than 0.8 according to cTWAS.

TWRCI annotates both cis and trans-variants, so we examined the locations of the annotated variants relative to the TSS for each of the eight causal genes. Most of the variants lying on the same chromosome as the TSS fell within a one megabase distance from the TSS (Figure 4 (e) blue). However, 78% of the variants were located on different chromosomes. We thus compared the variants annotated to causal genes by TWRCI against a previously published list of trans-eQTLs associated with any phenotype in a large-scale search^29^ (Methods 4.7.3). Variants annotated by TWRCI were located 1.94 times closer to trans-eQTLs than expected by chance (10,000 permutations, *p* < 0.001, 95% CI [1.93,1.95]). We next examined the effect sizes of the variants that cause the phenotype. We regressed the phenotype on variants inferred to directly or indirectly cause the phenotype using linear ridge regression. We then computed the moving average of the magnitudes of the regression coefficients over different distances from the TSS. The magnitudes remained approximately constant with increasing distance from the TSS (Figure 4 (e) red). Moreover, the magnitudes for variants located on different chromosomes did not converge to zero (dotted line). We thus conclude that trans-variants play a significant role in modulating gene expression to cause COPD.

#### 2.3.3 Root Causal Mechanism

We next analyzed the output of TWRCI to elucidate the root causal mechanism of COPD. The pathogenesis of COPD starts with inhaled irritants that trigger an exaggerated and persistent activation of inflammatory cells such as macrophages, T cells and B cells^23^. These cells in turn regulate a variety of inflammatory mediators that promote alveolar wall destruction, abnormal tissue repair and mucous hypersecretion obstructing airflow. The root causal genes of COPD therefore likely involve genes mediating chronic and exaggerated inflammation in the lung.

Eight of the fourteen gene expression levels ultimately caused the COPD phenotype in the causal graph reconstructed by TWRCI (Figure 4 (f)). The graph contained five MHC class II genes that present extracellular peptide antigens to CD4+ T cells in the adaptive immune response^30^. Subsequent activation of T cell receptors regulates a variety of inflammatory mediators and cytokines^31^. Moreover, the complement fragment C4a^32^ as well as the psoriasis susceptibility genes PSORS1C1 and PSORS1C2^33^ help initiate and maintain the exaggerated inflammatory response seen in COPD. The recovered causal graph thus implicates chronic exaggerated inflammation as the root causal mechanism of COPD. TWRCI replicated these results by again discovering C4A and the MHC class II genes in an independent GWAS dataset composed of individuals of East Asian ancestry (Supplementary Figure 3 (a)).

We finally analyzed the personalized CRCE estimates in more detail. We can decompose the CRCE estimate of each gene into genetic and non-genetic components according to Equation (1). The genetic variants explained only 6.4% of the estimated variance of the CRCE for HLA-DRB5, 1.4% for C4A and <1% for the other six causal genes. We conclude that non-genetic factors account for nearly all of the explained variance in the CRCE estimates. We then performed UMAP dimensionality reduction^34^ on the causal gene expression levels. Hierarchical clustering with Ward’s method^35^ yielded three clear clusters of patients with COPD (Figure 4 (g)) according to the elbow method on the sum of squares plot (Supplementary Figure 2 (a)). UMAP differentiated two of the COPD clusters from healthy controls, each with different mean CRCE estimates (Figure 4 (h) directed graphs). For example, HLA-DRB5 had a large positive CRCE in cluster one but a large negative CRCE in cluster two. The CRCE estimates thus differentiated multiple subgroups of patients consistent with the known pathobiology of COPD; we likewise obtained similar results in the second GWAS dataset (Supplementary Figure 3 (b) and (c)).

### 2.4 Oxidative stress in ischemic heart disease

We also ran the algorithms on summary statistics of ischemic heart disease (IHD) consisting of 31,640 cases and 187,152 controls from Finland^36^. We used variant-expression-phenotype data of whole blood from GTEx^22^ with 113 cases and 547 controls. We used whole blood because IHD arises from narrowing or obstruction of the coronary arteries most commonly secondary to atherosclerosis with transcription products released into the bloodstream^37^. We replicated the results using an independent set of GWAS summary statistics from 20,857 cases and 340,337 controls from the UK Biobank^38^.

#### 2.4.1 Accuracy

We compared the algorithms in variant annotation, graph reconstruction and CRCE estimation accuracy. TWRCI achieved the lowest MACR in both cases (Figure 5 (a) and (b)) within about one hour (Supplementary Figure 4 (b) and (c)). Cis-eQTLs and colocalization with SuSIE failed to annotate many variants because many trans-variants again predicted gene expression. We obtained similar results with a set of silver standard genes downloaded from the KEGG database (hsa05417)^39^, where TWRCI outperformed all other algorithms (Figure 5 (c)).

**Figure 5.**
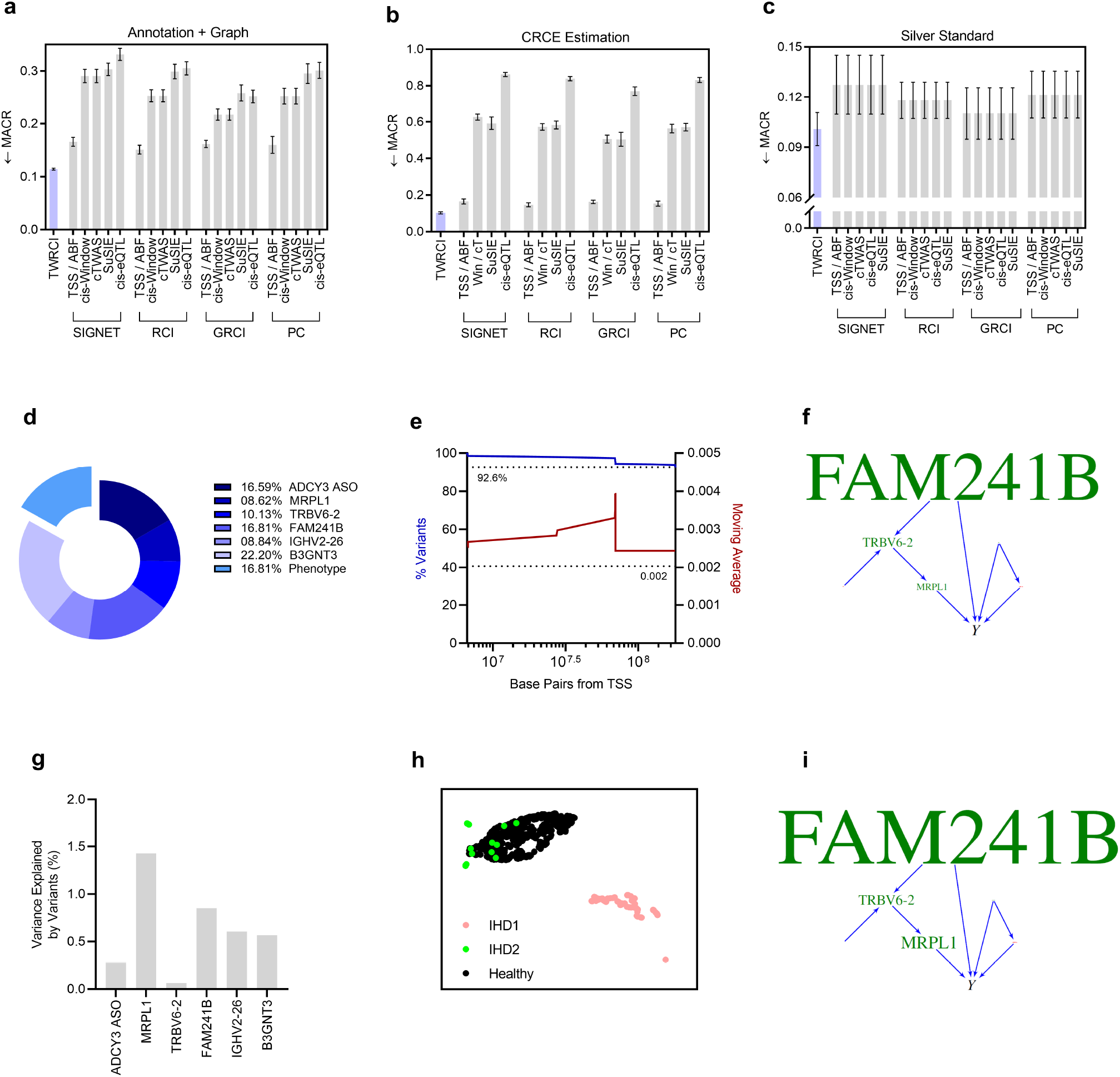
Results for IHD. (a) TWRCI again outperformed all other algorithms in combined annotation and graph reconstruction by achieving the lowest MACR. (b) TWRCI also estimated the CRCEs most accurately relative to all possible combinations of the other algorithms. (c) TWRCI outperformed all other algorithms with a silver standard set of genes causally involved in atherosclerosis. (d) TWRCI annotated variegated numbers of variants to six causal expression levels as well as the phenotype. (e) Nearly all of the annotated variants were located distal to the TSS (blue), and the magnitudes of their causal effects did not consistently increase or decrease on average with greater distance from the TSS (red). (f) TWRCI estimated the largest mean CRCEs for MRPL1, TRBV6-2 and FAM241B. (g) The annotated variants only explained a small proportion (<1.5%) of the variance for all CRCE estimates. (h) UMAP dimensionality reduction identified one cluster of patients clearly separated from healthy controls. (i) The mean CRCEs of MRPL1, TRBV6-2 and FAM241B remained the largest in this cluster.

#### 2.4.2 Horizontal pleiotropy and trans-variants

The genetic variants predicted 27 gene expression levels at an FDR threshold of 10% with six genes inferred to cause the phenotype. We plot the six genes in the directed graph recovered by TWRCI in Figure 5 (f). TWRCI sorted approximately 8-23% of the causal variants to each of the six genes (Figure 5 (d)). Moreover, TWRCI annotated approximately 17% of the causal variants directly to the phenotype supporting widespread horizontal pleiotropy in IHD. In contrast, cTWAS again did not detect any variants that directly cause the phenotype with a posterior inclusion probability greater than 0.8.

We analyzed the inferred causal effects of cis and trans-variants. Only 7.4% of the annotated variants were located on the same chromosome, and those on the same chromosome were often located over 10 megabases from the TSS (Figure 5 (e) blue). Moreover, variants annotated by TWRCI were located 4.46 times closer to a published list of trans-eQTLs than expected by chance (10,000 permutations, *p* = 0.0014, 95% CI [4.39,4.52]). The magnitudes of the regression coefficients remained approximately constant with increasing distance from the TSS and converged to 0.002 – rather than to zero – on different chromosomes (Figure 5 (e) red). We conclude that trans-variants also play a prominent role in IHD.

#### 2.4.3 Root Causal Mechanism

We next examined the root causal genes of IHD. IHD is usually caused by atherosclerosis, where sites of disturbed laminar flow and altered shear stress trap low-density lipoprotein (LDL)^40^. Reactive oxygen species then oxidize LDL and stimulate an inflammatory response. T cells in turn stimulate macrophages that ingest the oxidized LDL. The macrophages then develop into lipid-laden foam cells that form the initial fatty streak of an eventual atherosclerotic plaque. We therefore expect the root causal genes of IHD to involve oxidative stress and the inflammatory response.

TWRCI identified MRPL1, TRBV6-2 and FAM241B as the top three root causal genes (Figure 5 (f)). MRPL1 encodes a mitochondrial ribosomal protein that helps synthesize complex proteins involved in the respiratory chain^41^. Deficiency of MRPL1 can lead to increased oxidative stress. TRBV6-2 encodes a T-cell receptor beta variable involved in the inflammatory response and accumulation of T-cells in the atherosclerotic plaque^42^. Moreover, knocking out FAM241B induces the cytoplasmic buildup of large lysosome-derived vacuoles that generate foam cells^43^. We conclude that the root causal genes identified by TWRCI correspond to known genes involved in the pathogenesis of IHD. Finally, TWRCI rediscovered MRPL1 in a second independent GWAS dataset (Supplementary Figure 5 (a)).

We next dissected the CRCE estimates in detail. The annotated variants explained less than 1.5% of the CRCE variance for MRPL1, TRBV6-2 and FAM241B (Figure 5 (g)). Non-genetic factors therefore account for the vast majority of the CRCE variance. UMAP dimensionality reduction and then hierarchical cluster on the causal genes discovered by TWRCI revealed two clusters of IHD patients (Supplementary Figure 4 (a)). The largest of the two clusters lied distal to the cluster of healthy controls (Figure 5 (h)). Furthermore, the FAM241B, TRBV6-2 and MRPL1 genes retained the largest mean CRCEs in this cluster (Figure 5 (i)). TWRCI likewise replicated the large mean CRCE estimate for MRPL1 in the independent GWAS dataset (Supplementary Figure 5 (a) and (b)). We conclude that the CRCE estimates also identify genes that differentiate patient subgroups in IHD.

## 3 Discussion

We introduced the CRCE of a gene, a measure of the causal effect of the genetic and non-genetic factors that directly cause a gene expression level on a phenotype. We then created the TWRCI algorithm that estimates the CRCE of each gene after simultaneously annotating variants and reconstructing the causal graph for improved statistical power. TWRCI annotates, reconstructs and estimates more accurately than alternative algorithms across multiple semi-synthetic and real datasets. Applications of TWRCI to COPD and IHD revealed succinct sets of root causal genes consistent with the known pathogenesis of each disease, which we verified by replication. Furthermore, clustering delineated patient subgroups whose pathogeneses were dictated by different root causal genes.

Our experimental results highlight the importance of incorporating trans-variants in statistical analysis. TWRCI annotated many variants distal to the TSS of each gene. These trans-variants improved the ability of the algorithm to learn models of gene regulation consistent with the correlations in the data according to the MACR criteria. Moreover, variants annotated by TWRCI were located closer to the positions of a previously published list of trans-eQTLs than expected by chance^29^. In contrast, nearest TSS, cis-windows, cTWAS, cis-eQTLs and the colocalization methods all rely on cis-variants that did not overlap with many GWAS hits both in the COPD and IHD datasets. Most GWAS hits likely lie distal to the TSSs in disease due to natural selection against cis-variants with large causal effects on gene expression^44^. As a result, algorithms that depend solely on cis-variants can fail to detect a large proportion of variants that cause disease in practice.

TWRCI detected widespread horizontal pleiotropy accounting for 13-17% of the causal variants in both the COPD and IHD datasets. Previous studies have detected horizontal pleiotropy in around 20% of causal variants even after considering thousands of gene expression levels as well^28^. Moreover, many of the variants annotated to the phenotype by TWRCI correlated with gene expression (Supplementary Figures 2 (d) and 4 (d)). Accounting for widespread horizontal pleiotropy thus mitigates pervasive confounding between gene expression levels and the phenotype.

The cTWAS algorithm did not detect widespread pleiotropy in the real datasets. The algorithm also underperformed TWRCI in the semi-synthetic data, even when we restricted the analyses to variants that directly cause the phenotype. We obtained these results because cTWAS relies on the SuSIE algorithm to identify pleiotropic variants. However, pleiotropic variants usually exhibit weak causal relations to the phenotype, so most of these variants do not achieve a large posterior inclusion probability in practice. Algorithms that depend on *absolute* measures of certainty, such as posterior probabilities or p-values, miss many causal variants with weak causal effects in general. TWRCI therefore instead annotates variants by relying on *relative* certainty via a novel process called Competitive Regression, which we showed leads to more consistent causal models across multiple metrics.

We re-emphasize that TWRCI is the only algorithm that accurately recovers *root* causal genes *initiating* pathogenesis. Other methods such as colocalization and cTWAS identify causal genes *involved* in pathogenesis, regardless of whether the genes are root causal or not root causal. As a result, only TWRCI inferred a few genes with large CRCE magnitudes even in complex diseases. Moreover, genes with non-zero CRCE magnitudes explained away most of the causal effects of the non-root causal genes in the silver standards. Both of these results are consistent with the omnigenic root causal model, or the hypothesis that only a few root causal genes initiate the vast majority of pathology in each patient even in complex disease by affecting a very large number of downstream genes^1^.

Recall that the above root causal genes differ from driver genes and core genes. Root causal genes generalize driver genes by accounting for all of the factors that directly influence gene expression levels across all diseases, rather than just somatic mutations in cancer^3^. Accounting for both genetic and non-genetic factors is especially important when non-genetic factors explain the majority of the variance in the root causal effects, as we saw in COPD and IHD. Finally, root causal genes differ from core genes, or the gene expression levels that directly cause a phenotype, by focusing on the beginning rather than the end of pathogenesis^2^. Root causal genes may affect the expression levels of downstream genes so that many genes are differentially expressed between patients and healthy controls including many core genes. A few root causal genes can therefore increase the number of core genes.

TWRCI provably identifies root causal genes and attains high empirical accuracy, but the algorithm carries several limitations. The algorithm cannot accommodate cycles or directed graphs with different directed edges, even though cycles may exist and direct causal relations may differ between patient populations in practice^45^. TWRCI also estimates CRCE values at the patient-specific level, but the CRCEs may also vary between different cell types. Finally, the algorithm uses linear rather than non-linear models to quantify the causal effects of the variants on gene expression or the phenotype. Future work should therefore consider relaxing the single DAG constraint and accommodating non-linear relations. Future work will also focus on scaling the method to millions of genetic variants without feature selection.

In summary, we introduced an algorithm called TWRCI for accurate estimation and interpretation of the CRCE using personalized causal graphs. TWRCI empirically discovers only a few gene expression levels with large CRCE magnitudes even within different patient subgroups of complex disease in concordance with the omnigenic root causal model^46^. We conclude that TWRCI is a novel, accurate and disease agnostic procedure that couples variant annotation with graph reconstruction to identify root causal genes using observational data alone.

## Data Availability

All datasets analyzed in this study have been previously published and are publicly accessible as described in Methods.

https://github.com/ericstrobl/TWRCI

## Acknowledgements

Research reported in this manuscript was supported by (1) the National Human Genome Research Institute of the National Institutes of Health under award numbers R01HG011138 and R35HG010718, and (2) the National Institute of General Medical Sciences of the National Institutes of Health under award number R01GM140287.

## 4 Methods

### 4.1 Background on Causal Discovery

Causal discovery refers to the process of discovering causal relations from data. We let italicized letters such as *Z*_*i*_ denote a singleton random variable and bold italicized letters such as ***Z*** denote sets of random variables. Calligraphic letters such as 𝒵 refer to sets of sets.

We consider a set of *p endogenous variables* ***Z***. We represent a causal process over ***Z*** using a *structural equation model* (SEM) consisting of a series of deterministic functions:

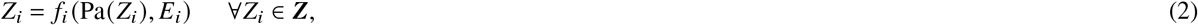

where Pa(*Z*_*i*_) ⊆ ***Z*** \ *Z*_*i*_ denotes the *parents*, of direct causes, of *Z*_*i*_ and *E*_*i*_ ∈ ***E*** an *exogenous variable*, also called an *error* or a *noise term*. We assume that the variables in ***E*** are mutually independent. The set Ch(*Z*_*i*_) refers to the *children*, or direct effects, of *Z*_*i*_ where *Z* _*j*_ ∈ Ch(*Z*_*i*_) if and only if *Z*_*i*_ ∈ Pa(*Z*_*j*_).

We can associate an SEM with a *directed graph* 𝔾 by a drawing a directed edge from *Z*_*j*_ to *Z*_*i*_ when *Z*_*j*_ ∈ Pa(*Z*_*i*_). We thus use the words *variable* and *vertex* interchangeably. A *root vertex* in 𝔾 refers to a vertex without any parents, whereas a *sink* or *terminal vertex* refers to a vertex without any children. A *path* between *Z*_0_ and *Z*_*n*_ corresponds to an ordered sequence of distinct vertices ⟨*Z*_0_, …, *Z*_*n*_⟩ such that *Z*_*i*_ and *Z*_*i*+1_ are adjacent for all 0 ≤ *i* ≤ *n* − 1. In contrast, a *directed path* from *Z*_0_ to *Z*_*n*_ corresponds to an ordered sequence of distinct vertices ⟨*Z*_0_, …, *Z*_*n*_⟩ such that *Z*_*i*_ ∈ Pa(*Z*_*i*+1_) for all 0 ≤ *i* ≤ *n* − 1. We say that *Z*_*j*_ is an *ancestor* of *Z*_*i*_, and likewise that *Z*_*i*_ is a *descendant* of *Z*_*j*_, if there exists a directed path from *Z*_*j*_ to *Z*_*i*_ (or *Z*_*j*_ = *Z*_*i*_). We collect all ancestors of *Z* _*j*_ into the set Anc(*Z*_*j*_), and all its non-descendants into the set Nd(*Z*_*j*_). We write *Z*_*i*_ ∈ Anc(***A***) when *Z*_*i*_ is an ancestor of any variable in ***A***, and likewise Nd(***A***) for the non-descendants. The variable *Z* _*j*_ *causes Z*_*i*_ if *Z*_*j*_ is an ancestor of *Z*_*i*_ and *Z*_*j*_ ≠ *Z*_*i*_. A *root cause* of *Z*_*i*_ corresponds to a root vertex that also causes *Z*_*i*_.

A *cycle* exists in 𝔾 when *Z*_*j*_ causes *Z*_*i*_ and vice versa. A *directed acyclic graph* (DAG) corresponds to a directed graph without cycles. A *collider* corresponds to *Z*_*j*_ in the triple *Z*_*i*_ → *Z*_*j*_ ← *Z*_*k*_. Two vertices *Z*_*i*_ and *Z*_*j*_ are d-connected given ***W*** ⊆ ***Z*** \ {*Z*_*i*_, *Z*_*j*_} if there exists a path between *Z*_*i*_ and *Z*_*j*_ such that no non-collider is in ***W*** and all colliders are ancestors of ***W***. We denote d-connection by 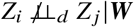 for shorthand. The two vertices are *d-separated* given ***W***, likewise denoted by *Z*_*i*_ ╨_*d*_ *Z* _*j*_ |***W***, if they are not d-connected. The *Markov boundary* of *Z*_*i*_, denoted by Mb(*Z*_*i*_), corresponds to the not necessarily unique but smallest set of variables in ***Z*** \ *Z*_*i*_ such that *Z*_*i*_ ╨_*d*_ (***Z*** \ Mb(*Z*_*i*_)) |Mb(*Z*_*i*_). A path is *blocked* by ***W*** if ***W*** contains at least one non-collider on the path or does not contain an ancestor of a collider (or both).

A probability density that obeys an SEM associated with the DAG 𝔾 also factorizes according to the graph:

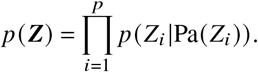

Any density that factorizes as above obeys the *global Markov property*, where *Z*_*i*_ and *Z*_*j*_ are conditionally independent given ***W***, or *Z*_*i*_ ╨ *Z* _*j*_ |***W***, if *Z*_*i*_ ╨_*d*_ *Z* _*j*_ |***W***^47^. A density obeys *d-separation faithfulness* when the converse holds: if *Z*_*i*_ ╨ *Z* _*j*_ |***W***, then *Z*_*i*_ ╨_*d*_ *Z*_*j*_ |***W***. The Markov boundary of *Z*_*i*_ uniquely corresponds to the parents, children and parents of the children (or *spouses*) of *Z*_*i*_ under d-separation faithfulness.

### 4.2 Causal Modeling of Variants, Gene Expression and the Phenotype

We divide the set of random variables ***Z*** into disjoint sets 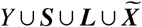 corresponding to the phenotype *Y, q* genetic variants ***S***, latent variables ***L*** modeling linkage disequilibrium (LD) and *m* gene expression levels 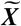. We model the causal process over ***Z*** using the following SEM associated with a DAG 𝔾:

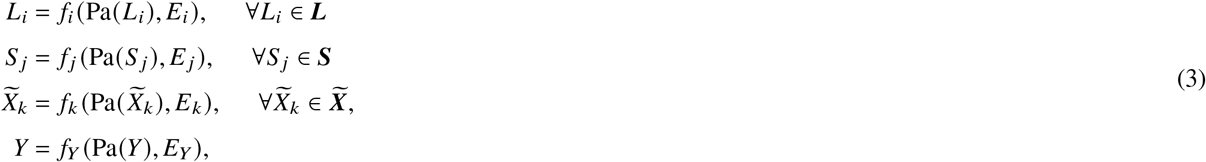

where 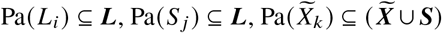 and 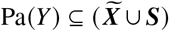 for any latent variable, any genetic variant, any gene expression level and the phenotype, respectively. In other words, linkage disequilibrium ***L*** generates variants ***S***, and variants and gene expression generate other gene expression levels 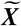 and the phenotype *Y* (example in Figure 6 (a)). We assume that *Y* is a sink vertex such that gene expression and variants cause *Y* but not vice versa.

**Figure 6.**
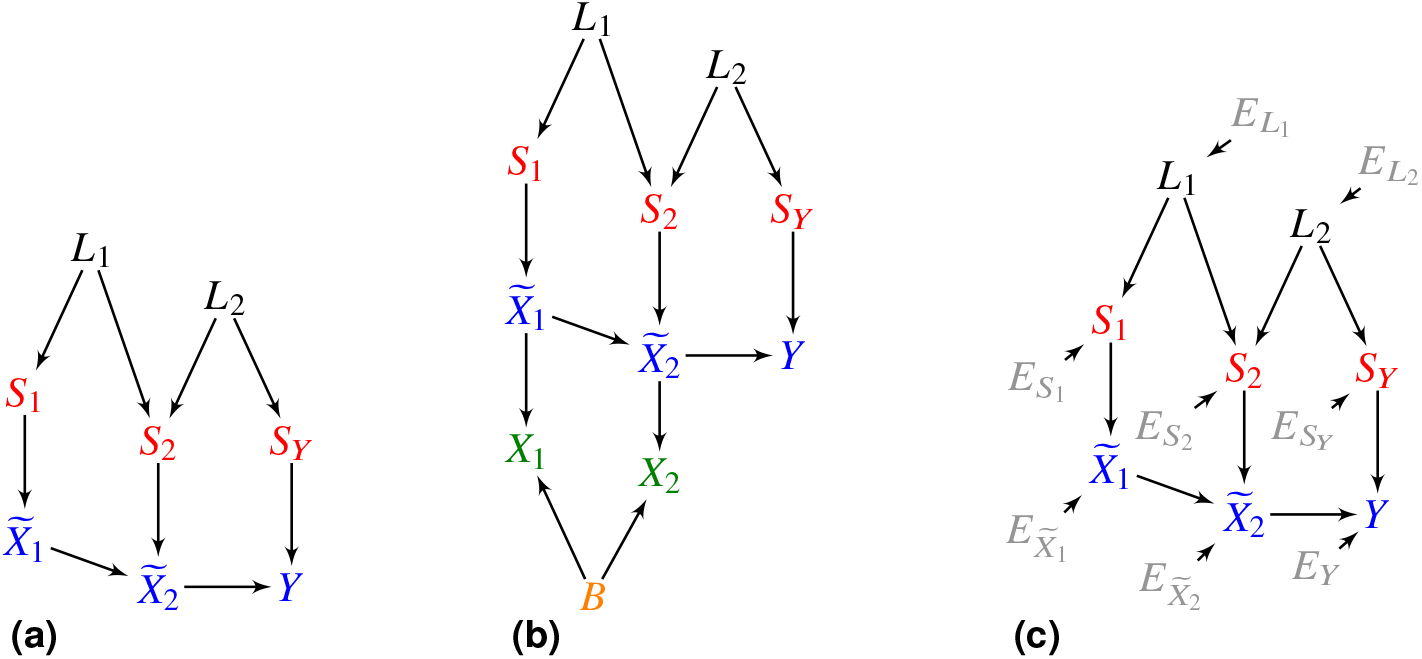
(a) An example of a DAG over ***Z***. In (b), the additional vertices ***X*** denote counts corrupted by batch *B* effects and Poisson measurement error. (c) We can also augment the DAG in (a) with root vertex error terms ***E***.

Let ***S***_*i*_ denote the direct causes of 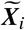 in ***S***. We require 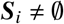 for all 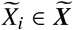 so that at least one variant directly causes each gene expression level. We also assume that any single variant can only *directly* cause one gene expression level or the phenotype (but not both). Investigators have reported only a few rare exceptions to this latter assumption in the literature^46^. A variant may however indirectly cause many gene expression levels.

We unfortunately cannot measure the exact values of gene expression using RNA sequencing (RNA-seq) technology. Numerous theoretical and experimental investigations have revealed that RNA-seq suffers from independent Poisson measurement error^48, 49^:

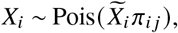

where *π*_*ij*_ denotes the *mapping efficiency* of 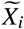 in batch *j*. We thus sample *Y* ⋃ ***S*** ⋃ ***L*** ⋃ ***X*** ⋃ *B* from the DAG like the one shown in Figure 6 (b) in practice, where *B* denotes the batch. With slight abuse of terminology, we will still call 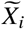 a *sink vertex* if it has only one child *X*_*i*_.

We can perform consistent regression under Poisson measurement error. Let 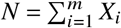 denote the library size and let 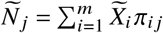 denote the true unobserved total gene expression level weighted by the mapping efficiencies in batch *j*. Also let 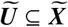 and ***V*** ⊆ ***S*** refer to any subset of gene expression levels and variants, respectively. The following result holds:

#### Lemma 1.

*Assume Lipschitz continuity of the conditional expectation for all N* ≥ *n*_0_:

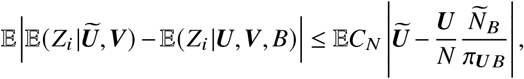

*where C*_*N*_ ∈ *O*(1) *is a positive constant, and we have taken an outer expectation on both sides. Then* 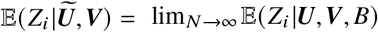 *almost surely*.

We delegate proofs to the Supplementary Materials. Intuitively, 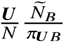 approaches 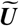 as the library size increases, so the above lemma states that accurate estimation of 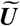 implies accurate estimation of 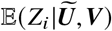. We can thus consistently estimate any conditional expectation 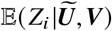 using 𝔼(*Z*_*i*_ | ***U, V, B***) when the library size approaches infinity. We only apply the asymptotic argument to bulk RNA-seq, where the library size is on the order of at least tens of millions. We henceforth implicitly assume additional conditioning on *B* whenever regressing to or on bulk RNA-seq data in order to simplify notation.

### 4.3 Conditional Root Causal Effects

We define the root causal effect of a gene expression level on the phenotype *Y*. We focus on Equation (3) with the endogenous variables ***Z*** and the exogenous variables ***E***. If the error terms ***E*** are mutually independent, then we can *augment* the associated DAG 𝔾 with ***E*** by drawing a directed edge from each 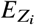 to its direct effect *Z*_*i*_ (Figure 6 (c)). We denote the resultant graph by 𝔾^′^, where we always have 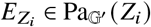 and the subscript emphasizes the augmented DAG; if we do not place a subscript, then we refer to the original DAG 𝔾. Only the error terms are root vertices in 𝔾^′^, so only exogenous variables that cause *Y* can be root causes of *Y*.

The *root causal effect* of *Z*_*i*_ on *Y* given the exogenous variables ***E*** is the causal effect of its direct causes in ***E*** on *Y*:

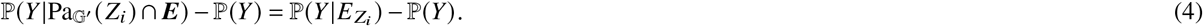

The variable *Z*_*i*_ is the first variable in ***Z*** affected by 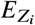, and *Z*_*i*_ may in turn causally affect *Y*. The exogenous variable 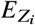 models the effects of environmental, epigenetic and other *non-genetic* factors on *Z*_*i*_ because the set of endogenous variables 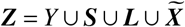 includes the *genetic factors* ***S***. The root causal effect is a special case of the *conditional root causal effect* (CRCE) given the exogenous variables ***E***:

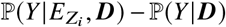

where 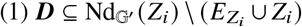 and 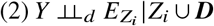. The first condition ensures that ***D*** does not block any directed path from *Z*_*i*_ to *Y*. The second ensures that ***D*** eliminates any confounding between 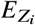 and *Y*. The first condition actually implies the second in this case because ***E*** are root vertices. If we set 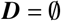, then we recover the unconditional root causal effect in Equation (4).

We are however interested in identifying the causal effects of both genetic *and* non-genetic factors on *Y* through gene expression 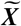 with potential confounding between members of ***S*** due to LD. We therefore expand the set of exogenous variables to ***E*** ⋃ ***S*** representing the non-genetic and genetic factors, respectively. We define the conditional root causal effect of 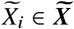 given the exogenous variables ***E*** ⋃ ***S*** as:

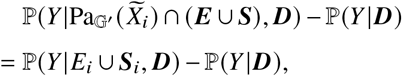

where we write 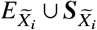 as *E*_*i*_ ⋃ ***S***_*i*_ to prevent cluttering of notation. The set *E*_*i*_ ⋃ ***S***_*i*_ thus refers to the direct causes of 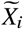 in ***E*** ⋃ ***S***. The above conditional root causal effect measures the causal effect of the root vertices ***E*** on *Y* as they pass through *E*_*i*_ ⋃ ***S***_*i*_ to 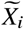.

We can likewise choose any ***D*** such that 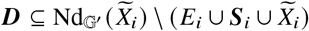 and 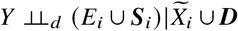. We choose ***D*** carefully to satisfy these two conditions as well as elicit favorable mathematical properties by setting 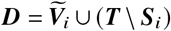, where 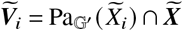 and 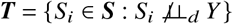. This particular choice of ***D*** allows us to write:

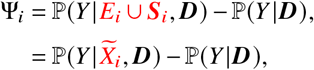

so that we do not need to recover *E*_*i*_ as an intermediate step. We prove the second equality in Proposition 1 of the Supplementary Materials under *exchangeability*, or no latent confounding by ***L*** between any two entries of 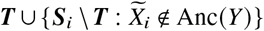; this union corresponds to a set of sets including ***T*** and each entry of 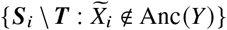 in the set. Exchangeability holds approximately in practice due to the weak causal relations emanating from variants to gene expression and the phenotype. Moreover, the assumption weakens with more variants in ***T***. Now the first gene expression level in 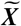 affected by *E*_*i*_ ⋃ ***S***_*i*_ is 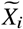. We thus call 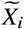 a *root causal gene* if 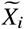 also causes *Y* such that Ψ_*i*_ ≠ 0.

We finally focus on the expected version of Ψ_*i*_ to enhance computational speed, improve statistical efficiency and overcome Poisson measurement error according to Lemma 1:

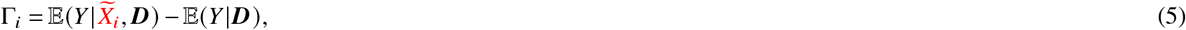

The *omnigenic root causal model* posits that |*γ*| ≫ 0 for only a small subset of gene expression levels in each patient with Γ = *γ*. We thus seek to estimate the values *γ* for each patient. We use the acronym CRCEs to specifically refer to Γ from here on.

### 4.4 Algorithm

#### 4.4.1 Strategy Overview

We seek to accurately annotate, reconstruct and estimate the CRCEs using (1) summary statistics as well as (2) linked variant-expression-phenotype data. We summarize the proposed Transcriptome-Wide Root Causal Inference (TWRCI) algorithm in Algorithm 1. TWRCI first uses summary statistics to identify variants ***T*** associated with the phenotype at a liberal *α* threshold in Line 1. The algorithm also identifies gene expression levels ***R*** ⊆ ***X*** predictable by ***T*** in Line 1 from the variant-expression-phenotype data. TWRCI then annotates non-overlapping sets of variants to the phenotype in Line 2 and each gene expression level in Line 3 using a novel process called Competitive Regression; we prove that annotated variants include all of the direct causes in ***T***. TWRCI identifies the causal ordering ***K*** among 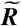 during the annotation process. The algorithm finally recovers the directed graph uniquely given ***K*** in Line 4 and estimates the CRCE of each gene inferred to cause *Y* using the estimated graph 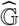 and the annotations 𝒫 in Line 5. TWRCI can thus weigh and color-code each node in 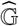 that causes *Y* by the CRCE estimates for each patient. We will formally prove that TWRCI is sound and complete at the end of this subsection.

##### Algorithm 1

Transcriptome-Wide Root Causal Inference (TWRCI)

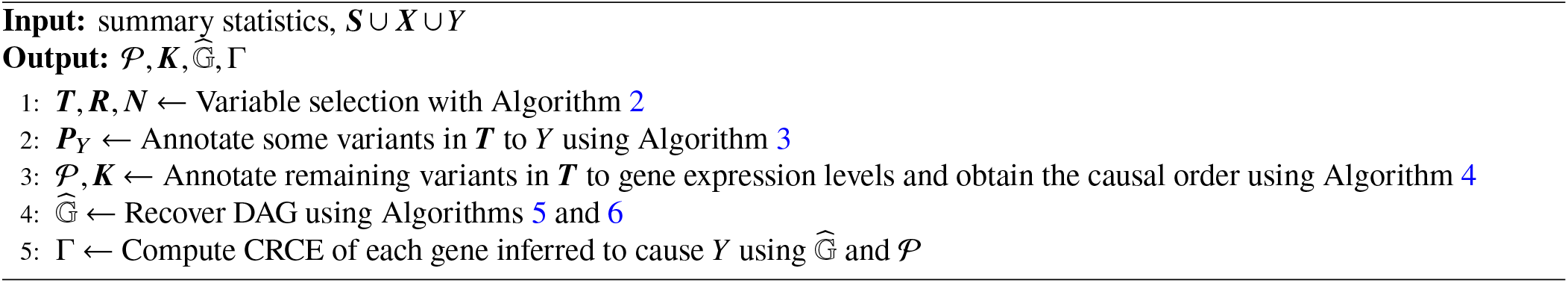

#### 4.4.2 Variable Selection

We summarize the variable selection portion of TWRCI in Algorithm 2. TWRCI first reduces the number of variants using summary statistics by only keeping variants with a significant association to the phenotype at a very liberal *α* threshold (Line 1); we use 5e-5, or a three orders of magnitude increase from the usual threshold of 5e-8. We do not employ clumping or other pre-processing methods that may remove more variants from consideration. Let ***T*** denote the variants that survive this screening step so that 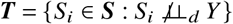.

The variable selection algorithm then identifies the gene expression levels predictable by ***T*** using the variant-expression-phenotype data in Line 2. We operationalize this step by linearly regressing ***X*** on ***T*** using half of the samples, and then testing whether the predicted level 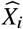 and the true level *X*_*i*_ linearly correlate in the second half for each *X*_*i*_ ∈ ***X***^50^. This sample splitting procedure ensures proper control of the Type I error rate^51^. We keep gene expression levels ***R*** ⊆ ***X*** that achieve a q-value below a liberal FDR threshold of 10%^52^. We say that ***T*** is *relevant* if it contains at least one variant that directly causes each member of 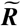. We finally repeat the above procedure after regressing out ***R*** from ***X*** \ ***R*** and ***T*** in Line 3 in order to identify *Ñ*, or all parents of 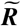 in 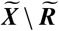. We call ***N*** the set of *nuisance variables*, since we will need to condition on them, but they do not contain the ancestors of *Y*. Algorithm 2 formally identifies the necessary ancestors needed for downstream inference:

##### Lemma 2.

*Assume d-separation faithfulness and relevance. Then*, 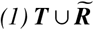 *contains all of the ancestors of Y in* 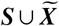, *and* 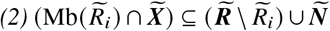 *for any* 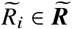.

##### Algorithm 2

Variable Selection

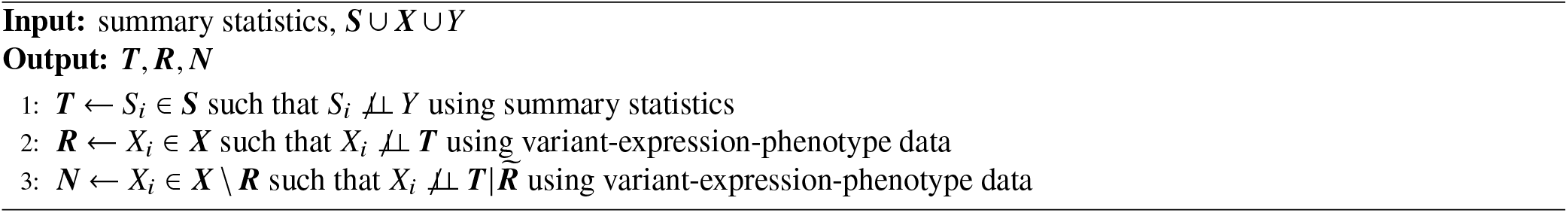

#### 4.4.3 Annotation for Horizontal Pleiotropy

TWRCI next annotates the associated variants ***T*** to their direct effects in 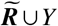. The algorithm first annotates a sink vertex and then gradually works its way up the DAG until it annotates the final root vertex.

TWRCI assumes that *Y* is a sink vertex, so it first annotates to *Y*. A variant exhibits *horizontal pleiotropy* if it directly causes *Y*. We propose a novel competitive regression scheme to annotate all members of ***T*** ⋂ Pa(*Y*) = ***S***_*Y*_ to *Y*.

We mildly assume equality in conditional expectation implies equality in conditional distribution and vice versa. Let 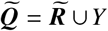 and likewise ***Q*** = ***R*** ⋃ *Y*. We also mildly assume that the following *contribution scores* exist and are finite: Δ_*ij*_ = 𝔼|*∂*𝔼(*Q*_*i*_ |***T***)/*∂T*_*j*_ | and 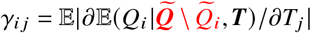. The scores correspond to the variable coefficients in linear regression. We use the contribution scores to annotate any *T*_*j*_ ∈ ***T*** such that 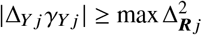 to *Y*, since this set of variants corresponds to a superset of ***S***_*Y*_ by the following result:

##### Corollary 1.

*Under d-separation faithfulness, relevance and exchangeability*, 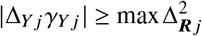 *if and only if T*_*j*_ ∉ Anc(***R***) *or T*_*j*_ ∈ Pa(*Y*) *(or both)*.

The proof follows directly from Lemma 3 in the Supplementary Materials.

The Competitive Regression (CR) algorithm summarized in Algorithm 3 computes the contribution scores in order to annotate variants to *Y*. Let Δ_−*i*_ denote the removal of the *i*^th^ row from Δ corresponding to *Q*_*i*_ = *Y*. We use debiased linear ridge regression^9^ to compute Δ in Line 1 and *γ*_*i*_ in Line 2. CR compares the two quantities and outputs the set 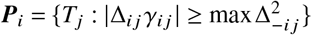, or a superset of ***S***_*i*_ ⋂***T*** not including any other variants with children in 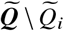 according to Corollary 1, in Line 3.

#### 4.4.4 Annotation and Causal Order

The CR algorithm requires the user to specify a known sink vertex. We drop this assumption by integrating CR into the Annotation and Causal Order (ACO) algorithm that automatically finds a sink vertex at each iteration.

ACO takes ***R, N***, *Y*, ***T, P***_*Y*_ as input as summarized in Algorithm 4. The algorithm constructs a causal ordering over ***R*** ⋃ *Y* in ***K*** by iteratively eliminating a sink vertex from ***R*** and appending it to the front of ***K***. ACO also instantiates a list 𝒫 and assigns genetic variants 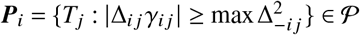 to each gene expression level *R*_*i*_ ∈ ***R*** in Lines 8 and 18 using the following generalization of Corollary 1:

##### Algorithm 3

Competitive Regression (CR)

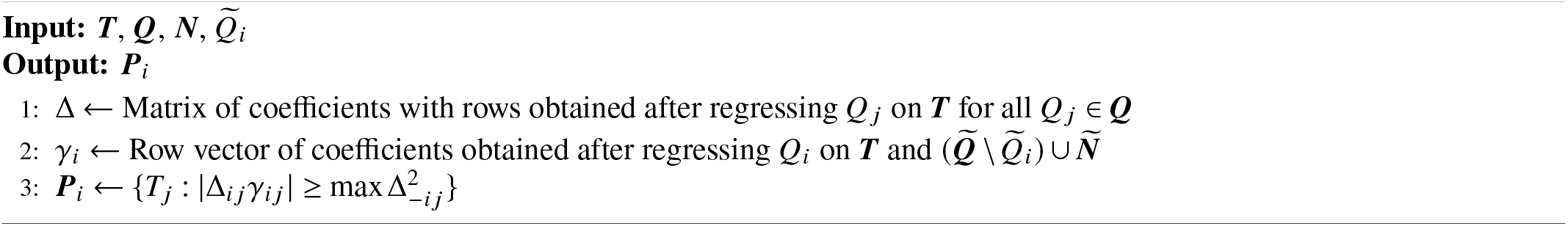

##### Algorithm 4

Annotation and Causal Order (ACO)

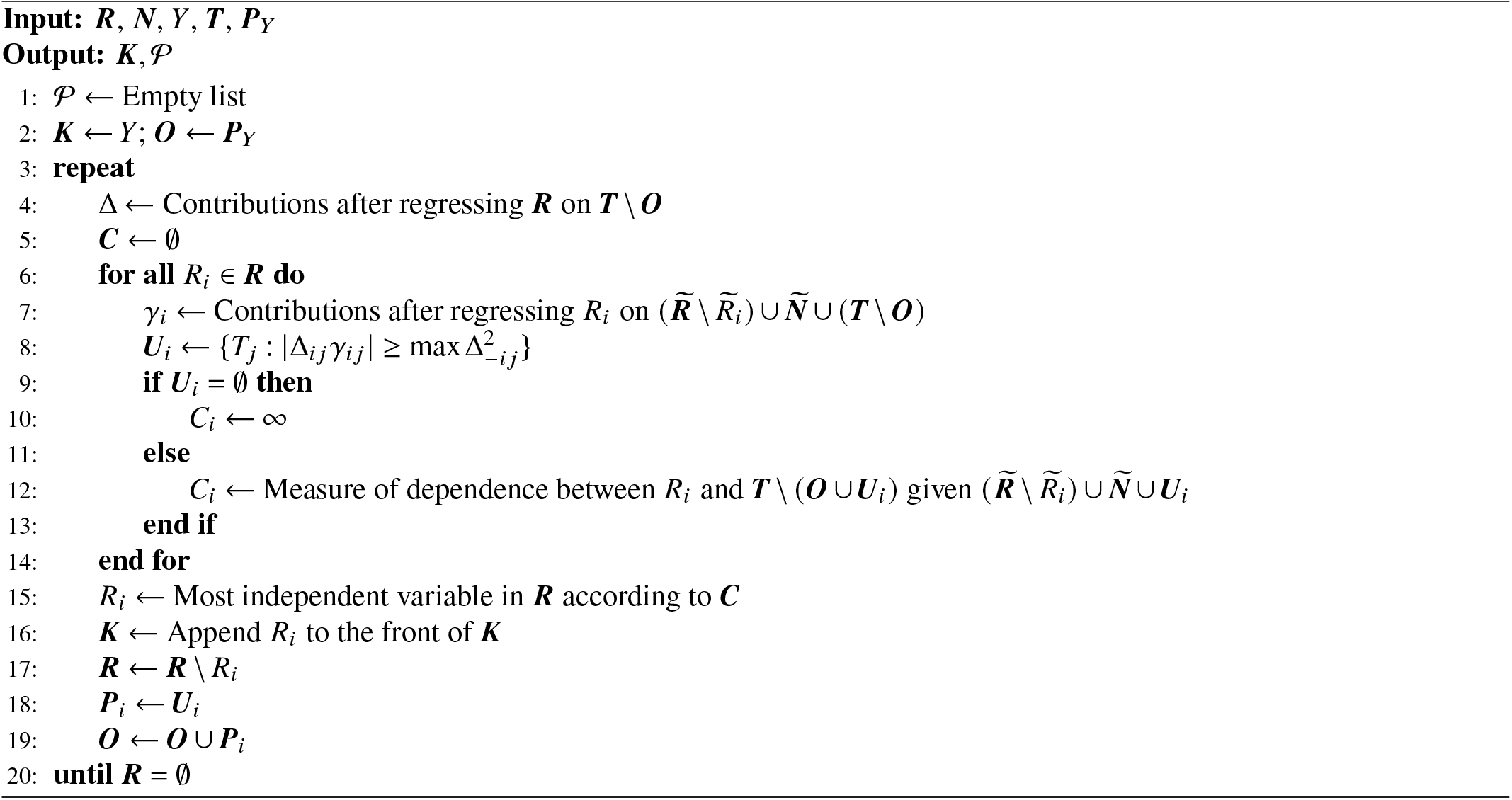

##### Lemma 3.

*Assume d-separation faithfulness, relevance and exchangeability. Further assume that* 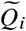 *is a sink vertex. Then*, 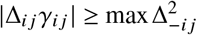 *if and only if T*_*j*_ ∉ Anc(***Q*** \ *Q*_*i*_) *or* 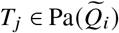 *(or both)*.

The set ***P***_*i*_ is thus again a superset of ***S***_*i*_ ⋂***T***, and any additional variants in ***P***_*i*_ do not directly cause another gene expression level or the phenotype.

ACO determines whether 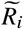 is indeed a sink vertex from data using the following result:

##### Lemma 4.

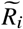 *is a sink vertex if and only if* 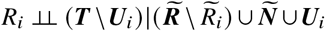 *in Line 12 of ACO under d-separation faithfulness, relevance and exchangeability*.

ACO practically determines whether any 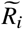 is indeed a sink vertex post variable elimination by first computing the residuals *F*_*i*_ after regressing *R*_*i*_ on 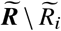, the nuisance variables *Ñ* and the identified variants ***U***_*i*_. A sink vertex 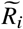 has residuals *F*_*i*_ that are uncorrelated with the variants in ***T*** \ (***O*** ⋃***U***_*i*_) in Line 12 by Lemma 4, so ACO can identify the sink vertex 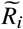 in Line 15 as the variable with the smallest absolute linear correlation. The algorithm then appends *R*_*i*_ to the front of ***K*** and eliminates *R*_*i*_ from ***R*** in Lines 16 and 17, respectively. ACO finally adds ***U***_*i*_ to 𝒫 in Line 18, so ***U***_*i*_ can be removed from ***T*** of the next iteration through ***O***. We formally prove the following result:

##### Lemma 5.

*Under d-separation faithfulness, relevance and exchangeability, ACO recovers the correct causal order* ***K*** *over* 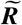 *and* (***S***_*i*_ ⋂***T***) ⊆ ***P***_*i*_ *for all* 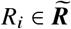.

#### 4.4.5 Causal Graph Discovery

TWRCI uses the causal order ***K*** and the annotations 𝒫 to perform causal discovery. The algorithm runs the (stabilized) skeleton discovery procedure of the Peter-Clark (PC) algorithm to identify the presence or absence of edges between any two gene expression levels (Algorithm 5)^19, 53^. We modify the PC algorithm so that it tests whether *R*_*i*_ and *R* _*j*_ are conditionally independent given ***P***_*i*_ ⋃ ***Ñ*** and subsets of the neighbors of 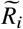 in 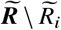 in Line 12 to ensure that we condition on all parents of 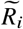. Finally, we orient the edges using the causal order ***K*** in Line 19 to uniquely recover the DAG over 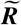:

##### Lemma 6.

*Under d-separation faithfulness, relevance and exchangeability, the graph discovery algorithm outputs the true sub-DAG over* 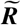 *given a conditional independence oracle*, ***K*** *and* 𝒫.

We next include the phenotype *Y* into the causal graph. We often only have a weak causal effect from gene expression and variants to the phenotype. We therefore choose to detect any causal relation to *Y* rather than just direct causal relations using Algorithm 6. Algorithm 6 only conditions on 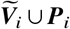 in Line 4 to discover both direct and indirect causation in concordance with the following result:

##### Lemma 7.

*Under d-separation faithfulness, relevance and exchangeability*, 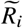 *causes Y – and likewise the vertices* ***S***_*i*_ ⋃ *E*_*i*_ *cause Y – if and only if* 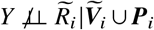.

##### Algorithm 5

Graph Discovery

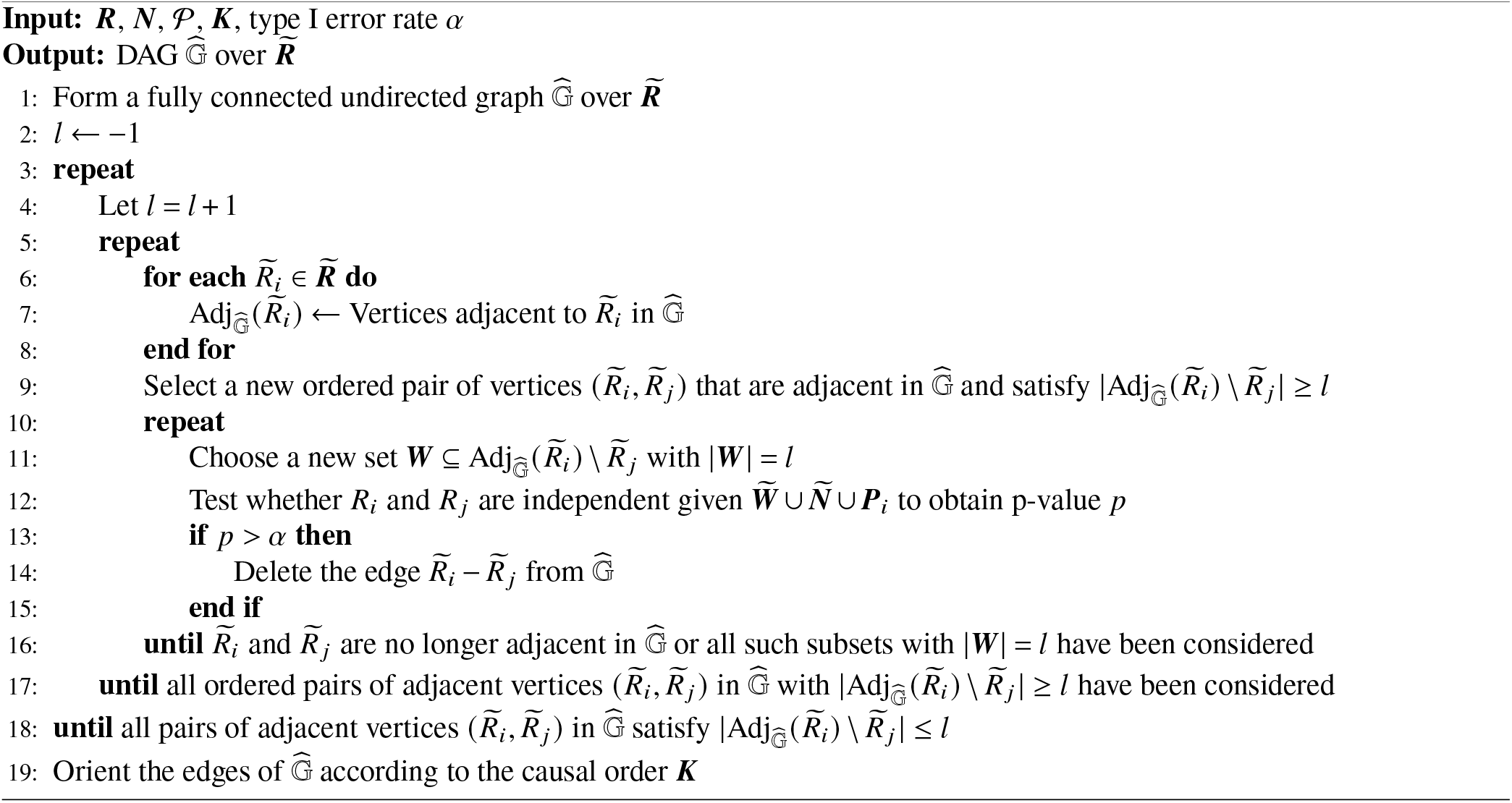

#### 4.4.6 Conditional Root Causal Effect Estimation

TWRCI finally estimates the CRCEs of the genes that cause *Y* given the recovered graph 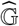 and the annotations 𝒫 We estimate the two conditional expectations in Equation (5) using kernel ridge regression^54^. We embed *X*_*i*_ and 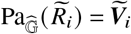 using a radial basis function kernel but embed ***T*** \ ***P***_*i*_ using a normalized linear kernel. We normalize the latter to prevent the linear kernel from dominating the radial basis function kernel, since the variables in ***T*** \ ***P***_*i*_ typically far outnumber those in 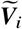.

We now integrate all steps of TWRCI by formally proving that TWRCI is sound and complete:

##### Algorithm 6

CRCE Graph Discovery

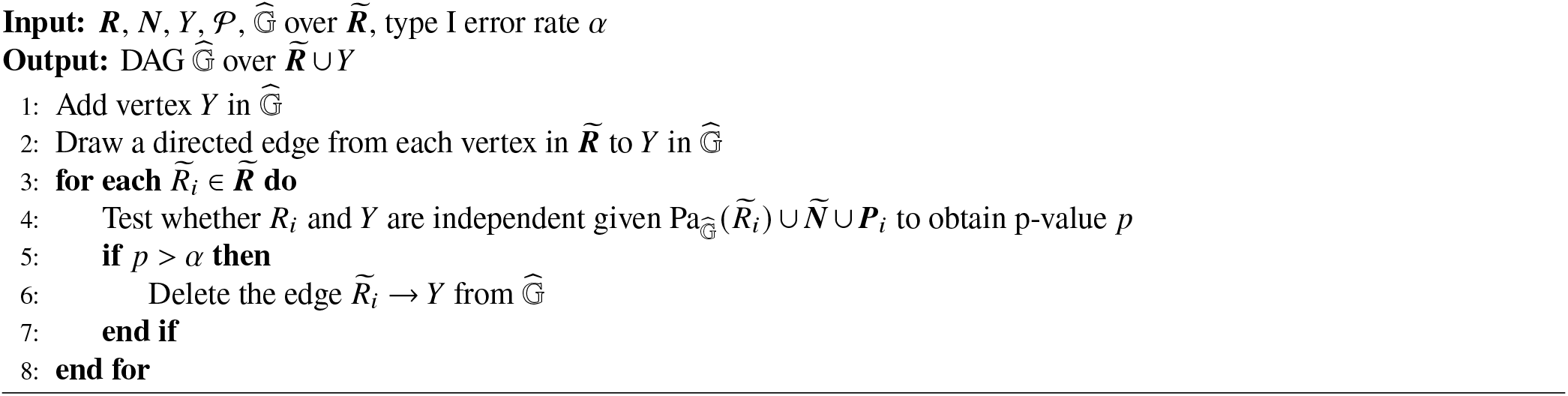

##### Theorem 1.

*(Fisher consistency) Under d-separation faithfulness, relevance and exchangeability, TWRCI identifies all of the direct causal variants of* 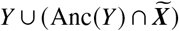, *the unique causal graph over* 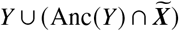 *and the CRCEs of* 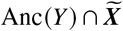 *almost surely as N* → ∞ *with Lipschitz continuous conditional expectations and a conditional independence oracle*.

We perform conditional independence testing by correlating the regression residuals of smooth non-linear transformations of the gene expression levels and phenotype^55^. As a result, Lemma 1 also enables accurate conditional independence testing over subsets of 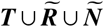, even though we only have access to ***T*** ⋃ ***R*** ⋃ ***N***.

#### 4.4.7 Time Complexity

We analyze the time complexity of TWRCI in detail. TWRCI can admit different regression procedures, so we will assume that each regression takes *O*(*c*^3^) time, where *c* denotes the dimensionality of the conditioning set typically much larger than the sample size *n*. Most regression procedures satisfy the requirement.

TWRCI first runs Algorithm 2 which requires *O*(*q*) time in Line 1 with summary statistics, *O*(*q*^3^*m*) time in Line 2 with at most *m* regressions on ***T***, and *O*(*m* (*m* + *q*)) time for at most *m* + *q* regressions on 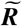 in Line 3. Algorithm 2 thus takes *O*(*m*^4^ + *m*^3^*q*) + *O*(*q*^3^*m*) time in total.

TWRCI next annotates to *Y* using Algorithm 3 which takes *O*(*q*^3^*m*) + *O*((*m* + *q*)^3^) time for Lines 1 and 2, respectively. Annotation to *Y* therefore carries a total time complexity of *O*(*m*^3^*q*^3^). TWRCI then runs Algorithm 4. Each iteration of the repeat loop in Line 3 of Algorithm 4 takes *O*(*q*^3^) time for the regression in Line 4 and *O*(*m*(*m* + *q*)^3^) time for the at most *m* regressions in Line 7. The repeat loop iterates at most *m* times, so Algorithm 4 has a total time complexity of *O*(*m*(*q*^3^ + *m*(*m* + *q*)^3^)) = *O*(*m*^5^*q*^3^).

Algorithm 5 dominates Algorithm 6 in time during the causal graph discovery portion of TWRCI. Algorithm 5 runs in *O*(*m*^*e*^ (*m* + *q*)^3^) = *O*(*m*^*e*+3^*q*^3^) time, where *e* denotes the maximum neighborhood size^19^. Finally, CRCE estimation in Line 5 requires *O*(2*m*(*m* + *q*)^3^) = *O*(*m*^4^*q*^3^) time for at most 2*m* regressions on expression levels and variants. Thus TWRCI in total requires *O*(*m*^4^ + *m*^3^*q*) + *O*(*q*^3^*m*) + *O*(*m*^3^*q*^3^) + *O*(*m*^5^*q*^3^) + *O*(*m*^*e*+3^*q*^3^) + *O*(*m*^4^*q*^3^) = *O*(*m*^5^*q*^3^) + *O*(*m*^*e*+3^*q*^3^) time. We conclude that the ACO and Graph Discovery sub-algorithms dominate the time complexity of TWRCI.

### 4.5 Comparators

We compared TWRCI against state of the art algorithms enumerated below.

#### Annotation

1. Nearest TSS: annotates each variant to its closest gene according to the TSS.
2. Cis-window: annotates a variant to a gene if the variant lies within a one megabase window of the TSS. If a variant lies in multiple windows, then we assign the variant to the closest TSS.
3. Causal transcriptome-wide association study (cTWAS)^11^: annotates variants to genes using cis-windows and then accounts for horizontal pleiotropy using the Sum of SIngle Effects (SuSIE) algorithm.
4. Cis-eQTLs^12^: annotates a variant to a gene if (1) the variant lies in the cis-window of the gene per above, and (2) the variant correlates most strongly with that gene expression level relative to the other levels.
5. Colocalization with approximate Bayes factors^13^: annotates each variant to the gene expression level with the highest colocalization probability according to approximate Bayes factors. We *could not* differentiate this method from cis-windows using the MACR criteria for the real data (Methods 4.8), since the algorithm always assigns higher approximate Bayes factors to cis-variants.
6. Colocalization with SuSIE^13, 14^: same as above but with probabilities determined according to SuSIE. We *could* differentiate this method from cis-windows using the MACR criteria for the real data.

#### Causal Graph Reconstruction

1. SIGNET^15, 16^: predicts gene expression levels from variants using ridge regression and then recovers the genetic ancestors of each expression level by running the adaptive LASSO on the predicted expression levels. The method thus assumes linearity.
2. RCI^17^: assumes a linear non-Gaussian acyclic model^56^, and recovers the causal order by maximizing independence between gene expression level residuals obtained from linear regression.
3. GRCI^18^: same as above but assumes an additive noise model^57^ and uses non-linear regression.
4. PC/CausalCell^20^: runs the stabilized PC algorithm^19, 53^ on the gene expression levels using a non-parametric conditional independence test^55^.

### 4.6 Semi-Synthetic Data

The causal graph reconstruction algorithms all require a variable selection step with gene expression data, since they cannot scale to the tens of thousands of genes with the neighborhood sizes seen in practice^1, 20^. We therefore assessed the performance of the algorithms independent of variable selection by first instantiating a DAG directly over 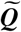 with *p* = 30 variables including 29 gene expression levels and a single phenotype. We generated a linear SEM obeying Equation (3) such that 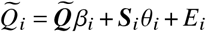 for every 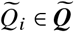 with *E*_*i*_ ∼ 𝒩 (0, 1/25) to enable detection of weak causal effects from variants. We drew the coefficient matrix *β* from a Bernoulli(2/(*p* − 1)) in the upper triangular portion of the matrix and then randomly permuted the ordering of the variables. The resultant DAG has an expected neighborhood size of 2. We then weighted the coefficient matrix between the gene expression levels and phenotype by sampling uniformly from [−1, −0.25] ∪ [0.25, 1].

We instantiated the variants ***T*** and *θ* as follows. We downloaded summary statistics from a wide variety of IEU datasets listed in Table 1 and filtered variants at a liberal *α* threshold of 5e-5. We selected a variant to be closest to the TSS of each gene uniformly at random and assigned direct causal variants to the 29 gene expression levels with probability proportional to the inverse of the absolute distance from the closest variant plus one. As a result, variants closer to the TSS are more likely to have a direct causal effect on the gene expression level. We assigned the remaining variants to the phenotype. We sampled ***T*** by bootstrap from the GTEx version 8^22^ individual-level genotype data and the weights *θ* uniformly from [−0.15, −0.05] ∪ [0.05, 0.15] because variants usually have weak causal effects.

**Table 1.**
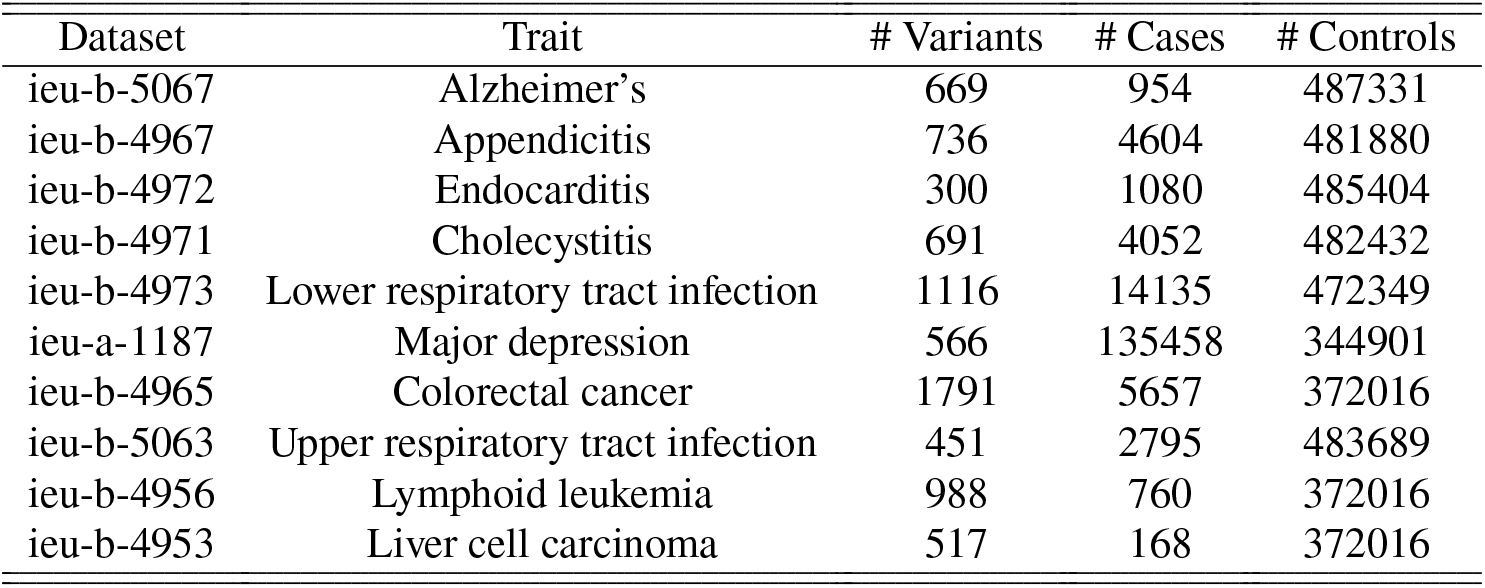
Variant data used during semi-synthetic data generation.

We converted the above linear SEM to a non-linear one by setting 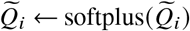 for each 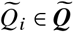. We obtained each measurement error corrupted surrogate *R*_*i*_ by sampling from 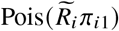 for each 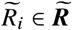. We drew the mapping efficiencies *π*_·1_ for a single batch from the uniform distribution between 100 and 10000 for the bulk RNA sequencing data. We repeated the entirety of the above procedure 100 times to generate 100 independent variant-expression-phenotype datasets. We ran TWRCI and all combinations of the comparator algorithms on each dataset.

### 4.7 Real Data

#### 4.7.1 Data Availability

All real datasets analyzed in this study have been previously published and are publicly accessible. The COPD datasets include:

1. Summary statistics: ebi-a-GCST90018807
2. Individual level variant and phenotype data: [GTEx V8 Protected Access Data](https://gtexportal.org/home/protectedDataAccess)
3. Gene expression data: [GTEx V8 Lung](https://storage.googleapis.com/adult-gtex/bulk-gex/v8/rna-seq/counts-by-tissue/gene_reads_2017-06-05_v8_lung.gct.gz)
4. Replication summary statistics: ebi-a-GCST90018587

The IHD datasets include:

1. Summary statistics: [finn-b-I9_ISCHHEART](https://gwas.mrcieu.ac.uk/datasets/finn-b-I9_IHD/)
2. Individual level variant and phenotype data: [GTEx V8 Protected Access Data](https://gtexportal.org/home/protectedDataAccess)
3. Gene expression data: [GTEx V8 Whole Blood](https://gtexportal.org/home/protectedDataAccess)
4. Replication summary statistics: ukb-d-I9_IHD

#### 4.7.2 Quality Control

We selected variants ***T*** at an *α* threshold of 5e-5 for both the COPD and IHD summary statistics. We harmonized the variant data of the IEU and GTEx datasets by lifting the GTEx variant data from the hg38 to hg19 build using the liftover command in BCFtools version 1.18^58^. We ensured that the reference and alternative alleles matched in both datasets after lifting for every variant. We removed gene expression levels with a mean count of less than five. We subjected the gene expression data to an inverse hyperbolic sine transformation to mitigate the effects of outliers. We regressed out the first 5 principal components, sequencing platform (Illumina HiSeq 2000 or HiSeq X), sequencing protocol (PCR-based or PCR-free) and sex from all variables in the linked GTEx variant-expression-phenotype data. Then, we either included age as a covariate for algorithms that accept a nuisance covariate, or regressed out age from the expression and phenotype data for algorithms that do not accept a nuisance covariate.

#### 4.7.3 Comparison to trans-eQTLs

TWRCI annotated many trans-variants in both of the real datasets. Other authors have proposed *trans-eQTLs* as variants that lie distal to the TSS and correlate with at least one reported phenotype in the Catalog of Published GWAS^59^. TWRCI annotates variants based on direct causality rather than correlation and an overlap with another phenotype. However, we hypothesized that the variants discovered by TWRCI should still lie close to at least a subset of the trans-eQTLs. To test this hypothesis, we downloaded trans-eQTL results from the [eQTLGen database](https://eqtlgen.org/trans-eqtls.html)^29^. We then standardized the positions of the variants within each chromosome by their standard deviation to account for variable chromosome length and polymorphism density. Next, we computed the nearest neighbor distances between the variants annotated to causal genes by TWRCI and the trans-eQTLs. We used the median of these normalized distances *M* as a robust statistic of central tendency.

We used a permutation test to test the null hypothesis that the variants annotated to causal genes by TWRCI are distributed arbitrarily far from the trans-eQTLs. We recomputed the median statistic 10,000 times after permuting the positions of the trans-eQTL variants. The p-value corresponds to the proportion of permuted statistics smaller than *M*. We reject the null hypothesis – and thus conclude that the variants annotated to causal genes by TWRCI lie close to trans-eQTLs – when the p-value falls below 0.05.

### 4.8 Metrics

We evaluated the accuracy of the algorithms using the nine metrics listed below for the synthetic data. We evaluated annotation quality using the following two metrics:

1. Matthew’s Correlation Coefficient (MCC)^60^ between the estimated annotations and the ground truth direct causal variants. Larger is better.
2. Rank of the estimated coefficients 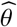 normalized by the rank of the ground truth coefficients *θ*. Larger is better. We also computed the above two quantities only using the variants that directly cause the phenotype in order to evaluate the ability of the algorithms to account for horizontal pleiotropy (3. and 4.). We evaluated the causal graph reconstruction quality using the following two metrics:
5. Structural Hamming Distance (SHD)^61^ between the estimated and the ground truth causal graph. Smaller is better.
6. MCC between the estimated and the ground truth causal graph. Larger is better. We evaluated combined annotation and graph reconstruction quality using Lemma 4:
7. Mean absolute correlation of the residuals (MACR) defined as the mean absolute correlation between (a) the variants ***T*** and ancestral gene expression levels, and (b) the gene expression residuals after partialing out the inferred parents. Smaller is better under the global Markov property and exchangeability. If the algorithm infers no direct causal variants in ***T*** and no parents in 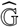 for some 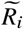, then this situation violates the relevance assumption, where at least one variant in ***T*** directly causes 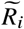. We thus set the absolute correlation of 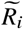 to one in this case. We assessed the accuracy in CRCE estimation using the following metrics:
8. Root mean squared error between the estimated CRCE and the ground truth CRCE averaged over all gene expression levels. We do not have access to the ground truth CRCE, so we estimate it to negligible error with kernel ridge regression using the ground truth parents. Smaller is better.
9. MACR between (a) the residuals 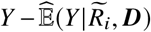 and (b) the inferred set ***P***_*i*_, which should be zero under the global Markov property and exchangeability. Smaller is better. We again set the absolute correlation to one for 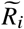 if the algorithm infers no direct causal variants and no parents in 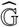 under relevance. We can compute the MACR metrics 7. and 9. on real data, so we evaluate the algorithms using these two metrics in the IHD and COPD datasets. We also have access to silver standard sets of genes known to be causally involved in disease from either the DisGeNet^26^ or KEGG database^39^. We therefore compute a third MACR metric with the real data:
10. A causal gene should at least correlate with the phenotype, so we first correlate the silver standard genes with the phenotype and only keep silver standard genes with a signification correlation (*p* < 0.05 uncorrected). We then compute a MACR metric between (a) the kept silver standard genes after partialing out genes with non-zero CRCEs and (b) the phenotype after partialing out genes with non-zero CRCEs.

### 4.9 Code Availability

R code needed to replicate all experimental results is available on GitHub.

## 5. Supplementary Materials

### 5.1 Additional Semi-Synthetic Data Results

**Supplementary Figure 1.**
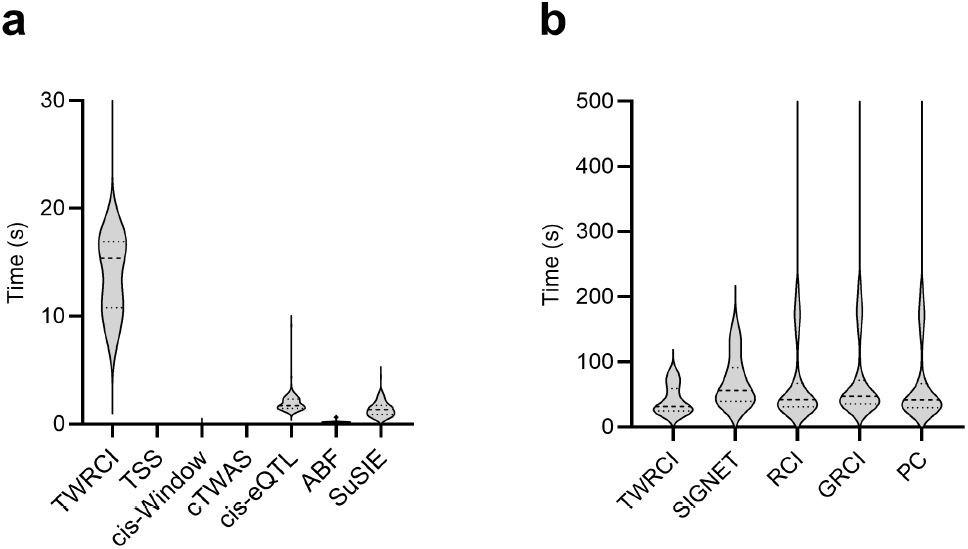
Timing results for the semi-synthetic datasets split into the variant annotation and graph reconstruction portions because they took the longest by far. (a) TWRCI took the longest time during annotation, but (b) all algorithms spent the majority of the time in causal graph reconstruction over 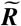 in congruence with the time complexity results of Methods 4.4.7. TWRCI completed within about 3 minutes overall.

### 5.2 Additional COPD Data Results

**Supplementary Figure 2.**
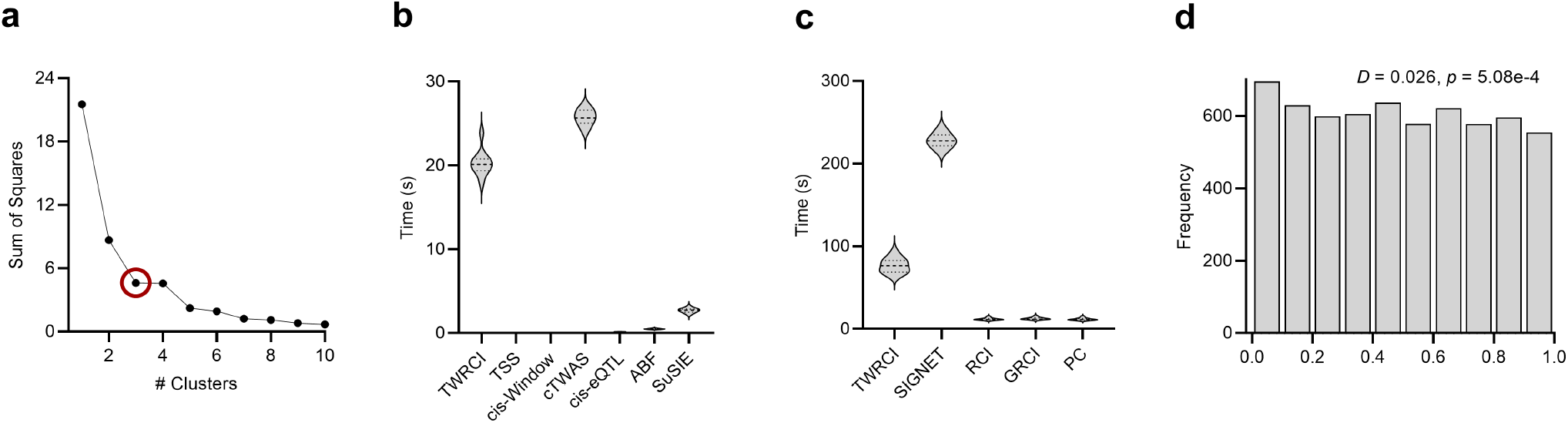
Additional results for COPD. (a) Sum of squares plot for hierarchical clustering using Ward’s method revealed three clusters according to the elbow method, or the cluster size with the maximum distance from the imaginary line drawn between the first and last cluster sizes. TWRCI took the second longest time to complete in annotation (b) and the second longest time to complete in graph reconstruction (c). RCI, GRCI and PC all took a much smaller amount of time to reconstruct the causal graph because they ignore the genetic variants. (d) Histogram of Pearson correlation test p-values computed between variants annotated to the phenotype and gene expression levels. The p-values did not follow a uniform distribution according to the Kolomogorov-Smirnov test with statistic *D* indicating the presence of confounding between the variants annotated to the phenotype and gene expression.

**Supplementary Figure 3.**
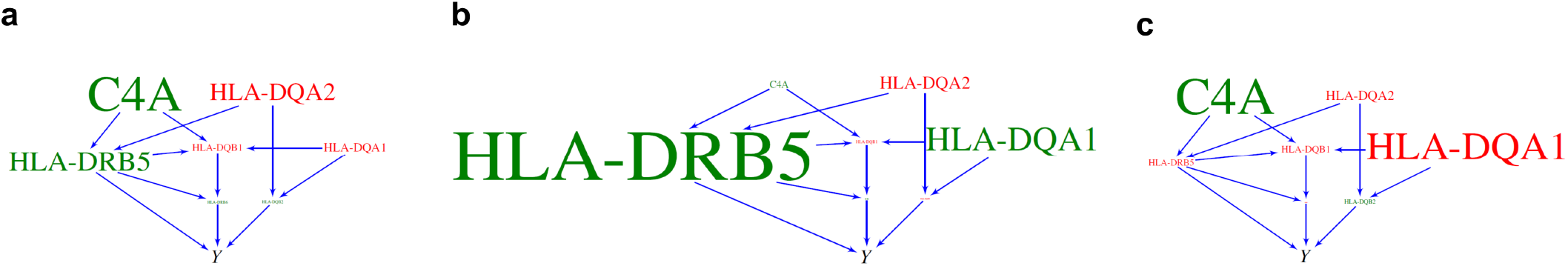
Replication results in an independent set of individuals of East Asian ancestry (dataset ebi-a-GCST90018587). We summarize results for (a) all patients, (b) cluster one in Figure 4 (g) and (c) cluster two. TWRCI again identified C4A and multiple MHC class II genes involving the adaptive immune system.

### 5.3 Additional IHD Data Results

**Supplementary Figure 4.**
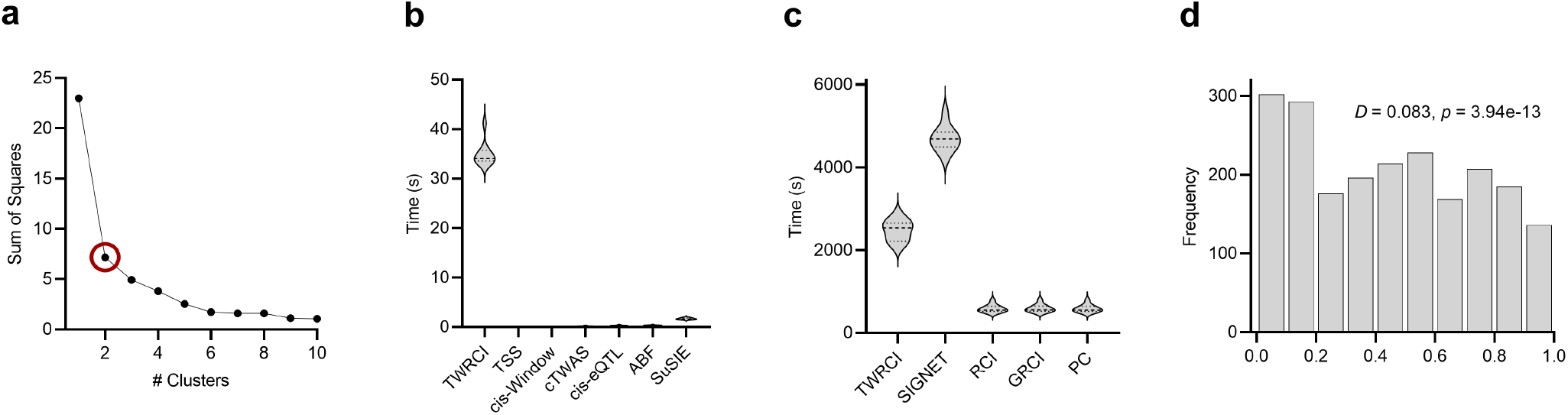
Additional results for IHD. (a) Sum of squares plot revealed two clusters according to the elbow method. (b) TWRCI took the longest to annotate, but (c) the timing results for graph reconstruction dominated in this case. Methods using SIGNET thus took the longest overall in this dataset. (d) Histogram of Pearson correlation test p-values were again non-uniform, indicating confounding between the variants annotated to the phenotype and gene expression.

**Supplementary Figure 5.**
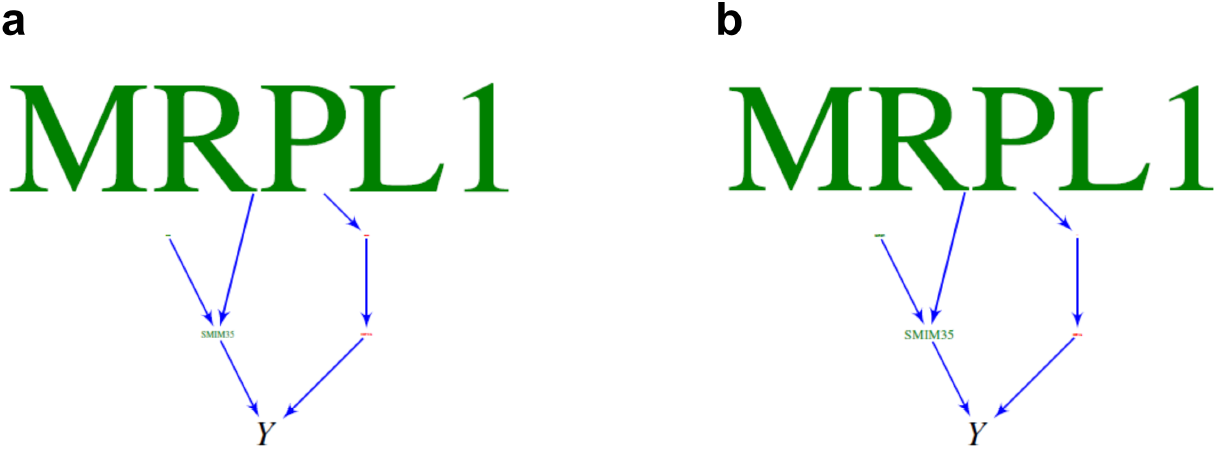
Replication results in an independent set of patients from the UK Biobank (dataset ukb-d-I9_IHD). We summarize results for (a) all patients, and (b) cluster one. TWRCI again identified MRPL1 as a root causal gene with a large positive CRCE.

### 5.4 Proofs

#### Lemma 1.

*Assume Lipschitz continuity of the conditional expectation for all N* ≥ *n*_0_:

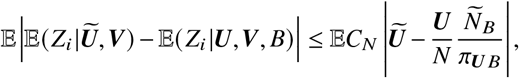

*where C*_*N*_ ∈ *O*(1) *is a positive constant, and we have taken an outer expectation on both sides. Then* 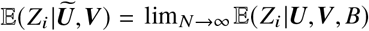 *almost surely*.

*Proof*. We can write the following sequence:

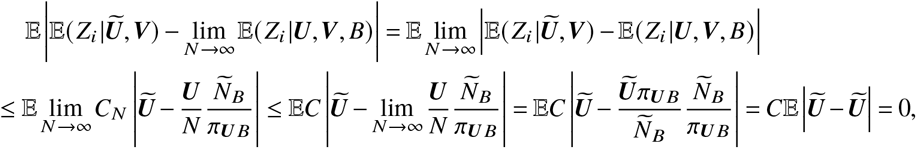

where we have applied the Lipschitz continuity assumption at the first inequality. We have *C*_*N*_ ≤ *C* for all *N* ≥ *n*_0_ in the second inequality because *C*_*N*_ ∈ *O*(1). With the above bound, choose *a* > 0 and invoke the Markov inequality:

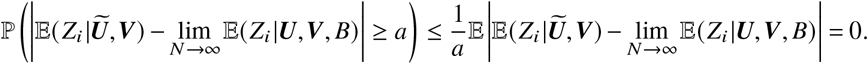

The conclusion follows because we chose *a* arbitrarily.

#### Proposition 1.

*We have* 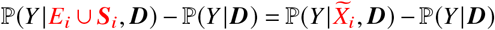 *under exchangeability. Proof*. We can write:

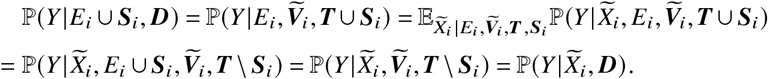

The third equality follows because 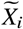 is a constant given *E* and 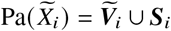. For the fourth equality, all paths between ***S***_*i*_ and *Y* are blocked by 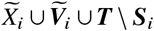 under exchangeability. We thus have 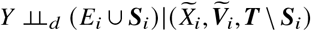 and 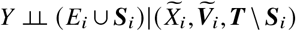 by the global Markov property.

#### Lemma 2.

*Assume d-separation faithfulness and relevance. Then*, 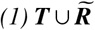 *contains all of the ancestors of Y in* 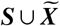, *and* 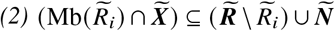 *for any* 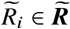.

*Proof*. We first prove (1). If *S*_*i*_ is an ancestor of *Y*, then 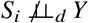, so 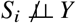 by d-separation faithfulness. It follows that *S* _*i*_ ∈ ***T*** by Line 1 of Algorithm 2. If 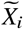 is an ancestor of *Y*, then so is ***S***_*i*_⊆ ***T***. Hence 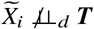,so 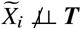 and 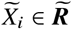 by d-separation faithfulness and Line 2, respectively. We chose *S*_*i*_ and 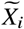 arbitrarily, so the set 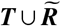 contains all of the ancestors of *Y* in 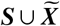.

We now prove (2). We need to show that 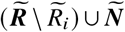 contains the parents, children and spouses of any 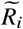, provided that these relatives are also in 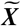. Note that 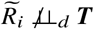 by Line 2 under the global Markov property. Hence, the parents and children of 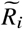 in 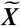 are also d-connected to ***T*** and hence dependent on ***T*** under d-separation faithfulness. It follows that 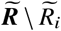 contains all of the parents and children of 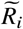 also by Line 2. Next, suppose 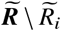 does not contain a spouse of 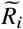, which we denote by 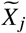. Then we have 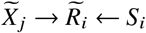 and *S*∈ ***T*** under relevance. Hence 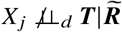, so 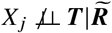 by d-separation faithfulness and *X*_*i*_∈***N*** by Line 3. It follows that 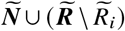 contains all of the spouses of 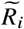. We conclude that 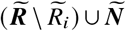 contains all members of 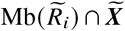 of any 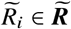.

#### Lemma 8.

*Under d-separation faithfulness, relevance and exchangeability, (1) T*_*j*_ ∉ Anc(*Q*_*i*_) *if and only if Q*_*i*_ ⫫ *T*_*j*_ |***T*** \ *T*_*j*_ *and* 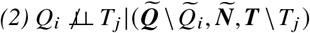 *and* 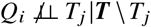 *if and only if* 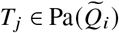.

*Proof*. For the first statement and forward direction, if *T*_*j*_ ∉ Anc(*Q*_*i*_), then *Q*_*i*_ ⫫_*d*_ *T*_*j*_ |***T*** \ *T*_*j*_ under exchangeability, so *Q*_*i*_ ⫫ *T*_*j*_ |***T*** \ *T*_*j*_ by the global Markov property. For the backward direction, if *Q*_*i*_ ⫫ *T*_*j*_ |***T*** \ *T*_*j*_, then *Q*_*i*_ ⫫_*d*_ *T*_*j*_ |***T*** \ *T*_*j*_ by d-separation faithfulness. No directed path can thus exist from *T*_*j*_ to *Q*_*i*_, so *T*_*j*_ ∉ Anc(*Q*_*i*_).

We next address the second statement. The backward direction follows immediately from d-separation faithfulness. For the forward direction, if 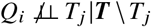, then *T*_*j*_ ∈Anc (*Q*_*i*_)from statement (1). Furthermore, if 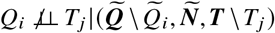 then 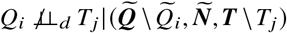 under the global Markov property. Note that 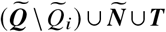 contains 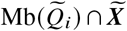 by Lemma 2 under d-separation faithfulness and relevance. Therefore, if *T*_*j*_ is not in the Markov boundary of 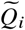, then 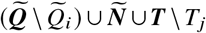 contains 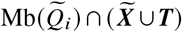. As a result, all paths between *T*_*j*_ and 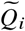 are blocked by 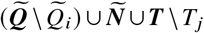 under exchangeability. We thus arrive at the contradiction 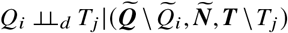. It follows that *T*_*j*_ must be in the Markov boundary of 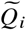 and therefore can only be a parent or a spouse of 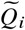 (or both). If *T*_*j*_ is a spouse but not a parent of 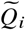, then we arrive at another contradiction that *T*_*j*_ ∉ Anc(*Q*_*i*_). Hence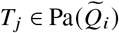.

#### Lemma 3.

*Assume d-separation faithfulness, relevance and exchangeability. Further assume that* 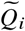 *is a sink vertex. Then*, 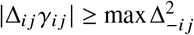 *if and only if T*_*j*_ ∉ Anc(***Q*** \ *Q*_*i*_) *or* 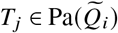*(or both)*.

*Proof*. Assume 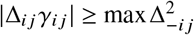 for the forward direction. We have two cases. If |Δ_*i j*_ *γ*_*i j*_ | > 0, then 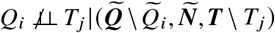, and 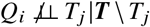, so 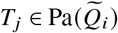 by Lemma 8. If |Δ_*i j*_ *γ*_*i j*_ | = 0, then 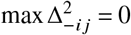, so *Q*_*k*_ ⫫ *T*_*j*_ |***T*** \ *T*_*j*_ for all *Q*_*k*_ ∈ ***Q*** \ *Q*_*i*_. We conclude that *T*_*j*_ ∉ Anc(***Q*** \ *Q*_*i*_) by again invoking Lemma 8.

For the backward direction, if *T*_*j*_ ∉ Anc(***Q*** \ *Q*_*i*_), then *Q*_*k*_ ⫫ *T*_*j*_ |***T*** \ *T*_*j*_ for all *Q*_*k*_ ∈ ***Q*** \ *Q*_*i*_ by Lemma 8. Thus 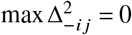 so 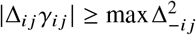. If 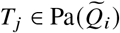, then *T*_*j*_ ∉ Anc (***Q*** \ *Q*_*i*_) because 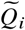 is a sink vertex. Hence *Q*_*k*_ ⫫ *T*_*j*_ |***T*** \ *T*_*j*_ for all *Q*_*k*_ ∈ ***Q*** \ *Q*_*i*_ by Lemma 8, so 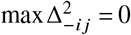. We conclude that 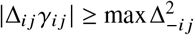.

#### Lemma 4.

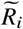 *is a sink vertex if and only if* 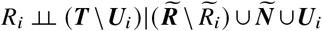 *in Line 12 of ACO under d-separation faithfulness, relevance and exchangeability*.

*Proof*. Assume that 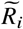 is a sink vertex for the forward direction. We have two cases:

1. If 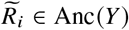, then 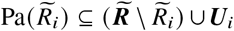 by the first statement of Lemma 2 and Lemma 3. Note that *Ñ* Hence, 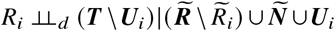 because 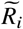 is a sink vertex, and 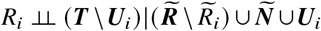 in Line 12 by the global Markov property.
2. If 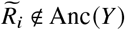, then 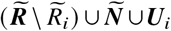 contains all of the parents of 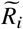 in 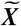 and ***T*** by Lemma 2 and Lemma 3, respectively. Moreover, the other direct causal variants of 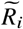, or ***S***_*i*_ \ ***T***, share no latent confounders with ***T*** or any other direct causal variant set excluding ***T*** by exchangeability. Hence, we also have 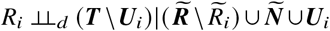, and 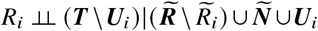 in Line 12 by the global Markov property.

We have exhausted all possibilities and thus conclude that 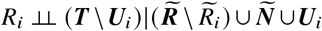.

For the backward direction, assume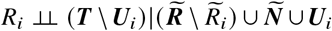 so that 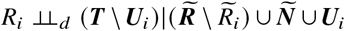 by d-separation faithfulness. Assume for a contradiction that 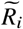 is not a sink vertex. Then there exists a path 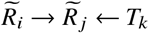 for some *T*_*k*_ ∈ ***S*** _*j*_ ∩***T*** by relevance. We thus have Δ*ik* = 0 but 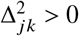, so *T*_*k*_ ∉ ***U***_*i*_ and *T*_*k*_ ∈ ***T*** \ ***U***_*i*_. We arrive at the contradiction 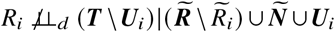 The variable 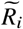 must therefore be a sink vertex.

#### Lemma 5.

*Under d-separation faithfulness, relevance and exchangeability, ACO recovers the correct causal order* ***K*** *over* 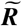 *and* (***S***_*i*_ ∩***T***) ⊆ ***P***_*i*_ *for all* 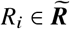.

*Proof*. We use proof by induction. <u>Base:</u> Suppose 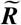 contains one variable 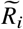. Then ***K*** = (*R*_*i*_,*Y*) because *R*_*i*_ is trivially the most independent variable in ***R*** according to ***C*** of Line 15. The variable *R*_*i*_ is a sink vertex after *Y* is eliminated, so we have (***S***_*i*_ ∩***T***) ⊆ ***P***_*i*_ under d-separation faithfulness, relevance and exchangeability by Lemma 3. Step: Assume that the conclusion holds when 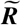 contains *p* − 1 variables. We need to prove the statement when 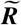 contains *p* variables. Assume for now that 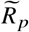 is an arbitrary sink vertex in 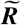. Lemma 3 then guarantees 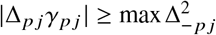 for each *S* _*j*_ ∈ ***S*** _*p*_ ∩ ***T*** in Line 8 under d-separation faithfulness, relevance and exchangeability. We thus have (***S*** _*p*_ ∩***T***) ⊆ ***P***_*p*_ and no variant of any other parent set is in ***P***_*p*_. Finally, the measure of dependence *C*_*p*_ in Line 15 identifies *R*_*p*_ as a sink vertex by Lemma 4. ACO thus eliminates *R*_*p*_ from ***R*** and appends it to the front of ***K***. The conclusion follows by the inductive hypothesis.

#### Lemma 6.

*Under d-separation faithfulness, relevance and exchangeability, the graph discovery algorithm outputs the true sub-DAG over* 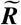 *given a conditional independence oracle*, ***K*** *and* 𝒫.

*Proof*. The set 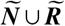 contains all of the parents of any 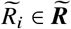 in 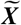 by Lemma 2. Furthermore, ***P***_*i*_ contains all of the parents of 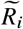 in ***T*** for any 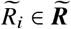 by Lemma 4. The stabilized skeleton discovery procedure of the PC algorithm thus recovers all and only the undirected edges in the true DAG over 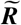 under d-separation faithfulness and exchangeability^53^. The conclusion follows because ACO recovers the true causal order over 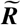 also by Lemma 5, so Algorithm 5 infers the true sub-DAG uniquely over 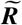 in Line 19.

#### Lemma 7.

*Under d-separation faithfulness, relevance and exchangeability*, 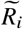 *causes Y – and likewise the vertices* ***S***_*i*_∪ *E*_*i*_ *cause Y – if and only if* 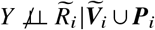.

*Proof*. Recall that (***S***_*i*_ ∩***T***) ⊆ ***P***_*i*_ by Lemma 5 under d-separation faithfulness, relevance and exchangeability.

Now if 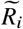 causes *Y*, then there exists a directed path from 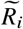 to *Y* so 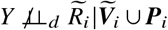. We then have 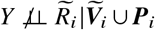 by d-separation faithfulness.

For the backward direction, assume that 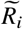 does not cause *Y*. All paths between 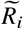 and *Y* are blocked by 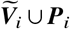 under exchangeability. Thus 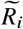 and *Y* are d-separated given 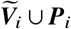. We invoke the global Markov property to conclude that 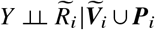.

#### Theorem 1.

*(Fisher consistency) Under d-separation faithfulness, relevance and exchangeability, TWRCI identifies all of the direct causal variants of* 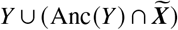, *the unique causal graph over* 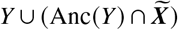 *and the CRCEs of* 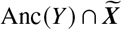 *almost surely as N* → ∞ *with Lipschitz continuous conditional expectations and a conditional independence oracle*.

*Proof*. Lemma 2 ensures 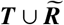 from Line 1 of Algorithm 1 contains all of the ancestors of *Y* in 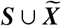. Thus 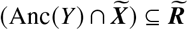 and (Anc (*Y*) ∩ ***S***⊆ ***T***). TWRCI identifies ***S***_*Y*_⊆ ***P***_*Y*_ in Line 2 by Lemma 3 under d-separation faithfulness, relevance and exchangeability. The algorithm also identifies ***S***_*i*_⊆ ***P***_*i*_ for each 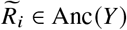 under d-separation faithfulness, relevance and exchangeability in Line 3 by invoking Lemma 5. Furthermore, TWRCI recovers the causal order over 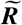 via Lemma 5. TWRCI thus uniquely recovers the sub-DAG over 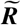 in Line 4 by Lemma 6 and then correctly includes *Y* in the graph by Lemma 7. TWRCI finally identifies the CRCEs of 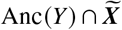 almost surely in Line 5 given the recovered DAG over 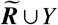 and 𝒫 by Lemma 1.

## Footnotes

1 If multiple genes were present in the window, then we assigned the variant to the gene with the nearest TSS.

